# Factors influencing self-harm thoughts and self-harm behaviours over the first year of the COVID-19 pandemic in the UK: a longitudinal analysis of 49,324 adults

**DOI:** 10.1101/2021.02.19.21252050

**Authors:** Elise Paul, Daisy Fancourt

**Affiliations:** Department of Behavioural Science and Health, University College London, 1-19 Torrington Place, London, WC1E 7HB, United Kingdom

**Keywords:** Self-harm behaviours, longitudinal studies, COVID-19, self-harm thoughts, adversity

## Abstract

**Background:** There is concern that the COVID-19 pandemic and its aftermath will result in excess suicides by increasing known risk factors such as self-harm, but evidence on how pandemic-related risk factors contribute to changes in these outcomes is lacking.

**Aims:** To examine how different COVID-19-related adverse experiences and adversity worries contribute to changes in self-harm thoughts and behaviours.

**Method:** Data from 49,324 UK adults in the UCL COVID-19 Social Study were analysed (1 April 2020 to 17 May 2021). Fixed effects regressions explored associations between weekly within-person variation in five categories of adversity experiences and adversity worries with changes in self-harm thoughts and behaviours across age groups (18-29, 30-44, 45-59, and 60+ years).

**Results:** 26.1% and 7.9% respondents reported self-harm thoughts and behaviours, respectively, at least once over the study period. More adverse experiences were more strongly related to outcomes than worries. The largest specific adversity contributing to increases in both outcomes was having experienced physical or psychological abuse. Financial worries increased the likelihood of both outcomes in most age groups, whilst having had COVID-19 increased the likelihood of both outcomes in young (18-29 years) and middle-aged (45-59 years) adults.

**Conclusions:** Findings suggest that a significant portion of UK adults may be at increased risk for self-harm thoughts and behaviours during the pandemic. Given the likelihood that the economic and social consequences of the pandemic will accumulate, policy makers can begin adapting evidence-based suicide prevention strategies and other social policies to help mitigate its consequences.

## Introduction

Numerous studies have shown that the COVID-19 pandemic is having a detrimental impact on population mental health, and although not inevitable, there are concerns that suicide rates will subsequently increase.^1^ Although other high-income countries have reported either no meaningful change or a decrease in suicide rates in the first months of the pandemic,^2^ temporarily lowered suicide rates have been observed in the early phases of other crises such as natural disasters and epidemics that were then followed by increases.^3^ This pattern has already been observed during the COVID-19 pandemic in Japan, where the first five months were marked by a 14% reduction in suicides, followed by a 16% increase in overall suicides, with a 49% increase in children and adolescents and a 37% increase in females.^4^ There are several reasons why suicides may increase once the immediate crisis has passed. First, the COVID-19 pandemic has involved the exacerbation of known risk factors for suicide such as unemployment, mental health problems, intimate partner violence, and insufficient access to mental health care that may not immediately resolve as the pandemic abates.^1,5^ Second, the cumulative effects of lockdowns, job losses, and uncertainty during the pandemic itself may start to take a toll over time.^1,5^ Third, the International Monetary Fund predicts that the global recession resulting from the COVID-19 pandemic will be the worst since the Great Depression,^6^ and research has consistently identified links between economic recessions, large scale unemployment, and increases in suicide rates.^7^ All of these stressful circumstances and life events have the potential to increase risk for suicide through increasing mental health difficulties such as depression, defeat, anxiety, and a sense of entrapment.^8,9^

One reason for concern about a potential future increase in suicide deaths as a result of the pandemic is that there is already evidence that risk factors for death by suicide have been increasing. Thinking about self-harming, suicide or death and intentionally damaging or injuring oneself have been widely observed to be risk factors for death by suicide.^10,11^ A number of studies have suggested that prevalence rates for thinking about self-harm or suicide or engaging in self-harming have been higher during the pandemic than previously.^12–14^ Although clinical presentations for self-harm have been significantly lower in the early months of the COVID-19 pandemic compared to prior trends,^15^ this could have been due to fears of contracting COVID-19 in hospitals and not wanting to be a burden on the healthcare system.^5,16^ Even pre-pandemic, the majority of individuals who self-harm or consider suicide do not seek help from clinical services.^17^

In considering why the above-mentioned risk factors (e.g., unemployment, mental health difficulties, and domestic abuse) for suicide may have increased in the first months of the current pandemic, several studies have identified potential predictors. Financial strain,^18^ experiencing physical/psychological abuse^12^ and receiving a COVID-19 diagnosis,^12,18^ legal problems, ongoing arguments with a partner, and worries about a life-threatening illness or injury in a family member or close friend have been associated with thinking about and carrying out self-harm,^19^ as have new and exacerbated mental health problems and insufficient access to mental health care.^20^ However, studies exploring predictors of self-harm thoughts and behaviours have been limited so far in which predictors they have considered and considering predictors at a single moment in time. As the social and economic circumstances of the pandemic are changing so fast, predictors identified early on in the pandemic may no longer be relevant. So, it is important to have updated information on what is causing people to think about harming themselves and to actually do so as the pandemic continues. Finally, it is important to identify which factors are associated over time not just with an increased overall risk in self-harm thoughts and behaviours but dynamic changes (both increases and decreases) so that modifiable targets to reduce self-harm can become the subject of future interventions.

Therefore, the aim of this study is to establish which factors are associated with changes over time in thoughts of death or self-harm (hereafter referred to as “self-harm thoughts”) and self-harm behaviours in a large sample of UK adults across the first 59 weeks of the COVID-19 pandemic. Specifically, we explore the time-varying longitudinal relationships between (i) adverse experiences and ii) worries about adverse experiences with changes in self-harm thoughts and behaviours, and how these associations vary by age. Identifying specific concerns and adversities that are risk factors for self-harm thoughts and behaviours will provide an opportunity for policy makers to address those issues by designing policies to mitigate the impact of the COVID-19 pandemic and the anticipated upcoming economic recession.

## Methods

### Study design and participants

Data were drawn from the COVID-19 Social Study; a large panel study of the psychological and social experiences of over 75,000 adults (aged 18+) in the UK during the COVID-19 pandemic. The study commenced on 21 March 2020 and involves online weekly data collection for the first 22 weeks of the COVID-19 lockdown in the UK (until 15 August 2020) then monthly collection thereafter. Sampling was not random and therefore is not representative of the UK population, but the sample is heterogeneous. More information on sampling methods can be found in the Supplementary Materials.

The authors assert that all procedures contributing to this work comply with the ethical standards of the relevant national and institutional committees on human experimentation and with the Helsinki Declaration of 1975, as revised in 2008. All procedures involving human subjects were approved by the UCL Ethics Committee. Written informed consent was obtained from all participants.

As questions asked about adverse experiences and worries about adversity in the prior week, we focused on data collected from 1 April 2020 (one week after lockdown commenced in the UK) to 17 May 2021 (n = 66,308; observations = 918,440). We then limited our analysis to participants who had taken part on three or more occasions during this period (n = 52,569; observations = 899,447). We further excluded participants who had missing data on any study variable for at least three interviews (n = 3,245). This resulted in the final sample of 49,324 participants totalling 849,452 observations (See Supplemental Table S1 for descriptive characteristics of excluded and included participants).

### Outcomes

#### Self-harm thoughts and behaviours

Self-harm thoughts were measured with an item from the Patient Health Questionnaire (PHQ-9)^21^ and self-harm behaviours were measured with a similar study-developed item (See Supplementary Materials). Responses to both items were collapsed into the presence (one or two days, more than half the days, or nearly every day) vs absence (not at all) at each time point.

### Exposures

#### Adversity experiences

Five categories of adversities measured weekly for the first 22 weeks of the study (1 April to 21 August 2020) and then monthly to 17 May 2021 were considered: financial adversity, COVID-19 illness, family/friend illness or bereavement, experiencing physical or psychological abuse, and not being able to access essential items. Each category of adversity was treated as binary (absent vs. present). More detailed description of these measures can be found in the Supplementary Materials.

#### Worries about adversity

Worries about adverse experiences were measured at the same time as the adversity measures and selected to correspond with these variables. Each category of worry was operationalised as binary (absent vs present): financial worries, COVID-19 illness, social and relationship worries, concerns about safety and security, and worries about accessing essentials. See the Supplementary Materials for further description of these measures.

#### Statistical Analysis

First, we describe weekly patterns in our outcome, adversity, and worries about adversity variables from 1 April 2020 through 17 May 2021. We then use fixed-effects regression to analyse the time-varying associations between changes in both experiences of and worries about adversity with changes in self-harm thoughts and behaviours across these 59 weeks. In this approach, individuals serve as their own reference point, which accounts for any confounding associations between time-invariant (stable) covariates such as socio-economic status, genetics, personality, and history of mental illness between predictors and outcomes.^22^ Our analyses consisted of regressing each outcome measure on i) the total number of adversity experiences and adversity worries jointly and ii) individual categories of adversity and worry about adversity for the total sample then stratified by age. All regression models adjusted for day of week (categorical) and days since lockdown commenced (continuous). Resulting regression coefficients were exponentiated and presented as odds ratios along with 95% confidence intervals. See Supplementary Materials for more detail, including model equation.

Sensitivity analyses included: models which included continuous measures of i) weekly depression symptoms, ii) weekly anxiety symptoms, and iii) the physical/psychological abuse variable separated into physical abuse and psychological abuse. To increase representativeness of the UK general population, data were weighted to the proportions of gender, age, ethnicity, country, and education in the UK, weights were constructed using the ‘ebalance’ programme in Stata^23^ based on data obtained from the Office for National Statistics.^24^ Analyses were conducted using Stata version 16.^25^

## Results

### Sample characteristics

In the unweighted analytic samples of participants with any change in self-harm thoughts (N = 11,580) or self-harm behaviours (N = 3,747) during the study period, women (78% in both samples) and individuals with a university degree or higher (69% in the self-harm thoughts sample; 64% in the self-harm behaviours sample) were overrepresented (Supplemental Table S2). In contrast, people from ethnic minority groups (6% in both samples) and young adults (ages 18-29; 10% self-harm thoughts; 14% self-harm behaviours) were underrepresented. After weighting, the samples reflected population proportions of these demographic characteristics (e.g., 55% women in both samples; people with a university degree or above: 36% in the self-harm thoughts sample; 30% in the self-harm behaviours sample; and 12% and 13% ethnic minorities in the self-harm thoughts and self-harm behaviours samples, respectively).

The average proportions of the sample reporting self-harm thoughts and self-harm behaviours over the first 59 weeks of the pandemic were relatively stable from the beginning of the pandemic to early autumn (Supplemental Figure S1a), but fluctuations were then seen in both outcomes in September and October 2020. The average number of total number of worries about adversity were consistently about three times higher than actual adversity experiences across the 59 weeks, with fluctuations seen starting in September 2020 (Supplemental Figure S1b), when data collection switched from weekly to monthly.

Over a quarter (26.1%) of respondents in the total sample reported having self-harm thoughts at least once over the first 59 weeks of the pandemic (Supplemental Table S3), and nearly one in ten (7.9%) had self-harmed at least once (Supplemental Table S4). There was within-individual variation over time in self-harm thoughts and behaviours outcome measures in 11,580 and 3,747 individuals, respectively, suggesting fixed effects was a valid approach. Descriptive statistics for predictor and outcome variables for each for these two samples are presented in Supplemental Table S5.

### Associations between total number of adversities and worries with self-harm thoughts and behaviours

Each additional adverse event experienced was associated with 1.56 (95% confidence interval [CI] = 1.52 to 1.60) times higher odds of self-harm thoughts (Supplemental Figure S2 & Table 1), whilst each additional adversity was associated with a nearly two-fold (odds ratio [OR] = 1.80; 95% CI = 1.72 to 1.87) increased likelihood of self-harm behaviours (Supplemental Figure S3 & Table 2) in the total sample. Increased likelihood of both outcomes in the total sample was smaller in magnitude for the total number of worries about adversity (self-harm thoughts: OR = 1.28; 95% CI = 1.26 to 1.30; self-harm behaviours: OR = 1.16; 95% CI = 1.13 to 1.19) than actual adversity experiences. This also applied to age-stratified analyses; the total number of adversity experiences (self-harm thoughts OR range = 1.43 to 1.61; self-harm behaviours OR range = 1.68 to 2.02) were more strongly associated with both outcomes than adversity worries in each of the four age groups (self-harm thoughts OR range = 1.19 to 1.33; self-harm behaviours OR range = 1.07 to 1.26).

**Table 1.** Fixed-effects logistic regression models predicting within-individual change in self-harm thoughts from the total number of adversity experiences and adversity worries

| Self-harm thoughts |  |  |  |  |  |  |  |  |  |  |  |  |  |  |  |
| --- | --- | --- | --- | --- | --- | --- | --- | --- | --- | --- | --- | --- | --- | --- | --- |
| Variable | Total sample |  |  | Ages 18-29 |  |  | Ages 30-44 |  |  | Ages 45-59 |  |  | Ages 60+ |  |  |
|  | OR | 95%<br>CI<br>lower | 95%<br>CI<br>upper | OR | 95%<br>CI<br>lower | 95% CI<br>upper | OR | 95% CI<br>lower | 95% CI<br>upper | OR | 95%<br>CI<br>lower | 95%<br>CI<br>upper | OR | 95%<br>CI<br>lower | 95%<br>CI<br>upper |
| Total number of<br>adversity experiences | 1.56 | 1.52 | 1.60 | 1.55 | 1.45 | 1.66 | 1.43 | 1.35 | 1.51 | 1.60 | 1.53 | 1.68 | 1.61 | 1.53 | 1.70 |
| Total number of<br>worries | 1.28 | 1.26 | 1.30 | 1.33 | 1.28 | 1.38 | 1.19 | 1.15 | 1.23 | 1.30 | 1.26 | 1.34 | 1.32 | 1.28 | 1.36 |
| Number of<br>observations | 206,714 |  |  | 16,495 |  |  | 53,752 |  |  | 74,635 |  |  | 61,832 |  |  |
| Number of individuals | 11,580 |  |  | 1,168 |  |  | 3,394 |  |  | 4,069 |  |  | 2,949 |  |  |

Note. Adversity experiences (0-5) and worries (0-5) variables are weighted to the proportions of gender, age, ethnicity, country, and education obtained from the Office for National Statistics. Individual adversity experiences and worries variables are binary. Analyses were further adjusted for day of the week and time since lockdown began. OR = odds ratio. CI = confidence interval.

**Table 2.** Fixed-effects logistic regression models predicting within-individual change in self-harm behaviours from the total number of adversity experiences and adversity worries

| Self-harm behaviours |  |  |  |  |  |  |  |  |  |  |  |  |  |  |  |
| --- | --- | --- | --- | --- | --- | --- | --- | --- | --- | --- | --- | --- | --- | --- | --- |
| Variable | Total sample |  |  | Ages 18-29 |  |  | Ages 30-44 |  |  | Ages 45-59 |  |  | Ages 60+ |  |  |
|  | OR | 95%<br>CI | 95%<br>CI | OR | 95%<br>CI | 95%<br>CI | OR | 95%<br>CI | 95%<br>CI | OR | 95%<br>CI | 95%<br>CI | OR | 95%<br>CI | 95%<br>CI |
|  |  | lower | upper |  | lower | upper |  | lower | upper |  | lower | upper |  | lower | upper |
| Total number of<br>adversity experiences | 1.80 | 1.72 | 1.87 | 1.68 | 1.54 | 1.83 | 1.70 | 1.55 | 1.86 | 1.79 | 1.67 | 1.93 | 2.02 | 1.85 | 2.21 |
| Total number of worries | 1.16 | 1.13 | 1.19 | 1.26 | 1.19 | 1.34 | 1.07 | 1.01 | 1.14 | 1.19 | 1.13 | 1.25 | 1.10 | 1.04 | 1.17 |
| Number of observations | 63,767 |  |  | 7,687 |  |  | 18,594 |  |  | 23,859 |  |  | 13,627 |  |  |
| Number of individuals | 3,747 |  |  | 530 |  |  | 1,201 |  |  | 1,329 |  |  | 687 |  |  |

Note. Adversity experiences (0-5) and worries (0-5) variables are weighted to the proportions of gender, age, ethnicity, country, and education obtained from the Office for National Statistics. Individual adversity experiences and worries variables are binary. Analyses were further adjusted for day of the week and time since lockdown began. OR = odds ratio. CI = confidence interval.

### Associations between individual categories of adversities and worries with self-harm thoughts and behaviours

When examining individual categories of adversities and adversity worries, having experienced psychological or physical abuse had the largest associations with both outcomes across all age groups and in the total sample (Supplemental Figures S4 and S5 and Tables 3 and 4). Odds ratios were slightly higher for self-harm behaviours (OR range: 3.39 to 6.96) than for self-harm thoughts (OR range: 3.37 to 3.90). Increases in financial adversities and worries, social/relationship concerns, and concerns about one’s safety (‘threats to safety’) increased the likelihood of later self-harm thoughts in all age groups. Financial concerns generally had larger magnitudes of association (self-harm thoughts OR range: 1.35 to 1.72; self-harm behaviours OR range: 1.11 to 1.40) with both outcomes than actual adversity experiences (self-harm thoughts OR range: 1.12 to 1.27; self-harm behaviours OR range: 0.80 to 1.14). Having had COVID-19 increased the likelihood of both outcomes in young adults and in adults ages 45-59. However, concerns about becoming ill with COVID-19 only increased the likelihood of self-harm thoughts, and only in older adults, and decreased likelihood of self-harm thoughts in the total sample, young adults, and adults ages 30-44.

**Table 3.** Fixed-effects logistic regression models predicting within-individual change in self-harm thoughts from individual categories of adversity experiences and worries

| Self-harm thoughts |  |  |  |  |  |  |  |  |  |  |  |  |  |  |  |
| --- | --- | --- | --- | --- | --- | --- | --- | --- | --- | --- | --- | --- | --- | --- | --- |
|  | Total sample |  |  | Ages 18-29 |  |  | Ages 30-44 |  |  | Ages 45-59 |  |  | Ages 60+ |  |  |
| Variable | OR | 95%<br>CI<br>lower | 95%<br>CI<br>upper | OR | 95%<br>CI<br>lower | 95%<br>CI<br>upper | OR | 95%<br>CI<br>lower | 95%<br>CI<br>upper | OR | 95%<br>CI<br>lower | 95%<br>CI<br>upper | OR | 95%<br>CI<br>lower | 95%<br>CI<br>upper |
| Adversity experiences |  |  |  |  |  |  |  |  |  |  |  |  |  |  |  |
| Financial | 1.18 | 1.13 | 1.25 | 1.27 | 1.12 | 1.42 | 1.16 | 1.05 | 1.29 | 1.12 | 1.03 | 1.22 | 1.25 | 1.12 | 1.40 |
| Accessing essentials | 1.28 | 1.19 | 1.37 | 0.93 | 0.76 | 1.14 | 1.00 | 0.87 | 1.17 | 1.66 | 1.46 | 1.89 | 1.33 | 1.17 | 1.52 |
| COVID-19 illness | 1.17 | 1.09 | 1.25 | 1.26 | 1.06 | 1.50 | 1.04 | 0.91 | 1.19 | 1.27 | 1.13 | 1.42 | 1.12 | 0.98 | 1.29 |
| Illness of others/bereavement | 1.14 | 1.08 | 1.21 | 1.29 | 1.10 | 1.50 | 1.02 | 0.89 | 1.16 | 1.23 | 1.11 | 1.37 | 1.09 | 0.98 | 1.21 |
| Physical/psychological abuse | 3.55 | 3.37 | 3.75 | 3.73 | 3.19 | 4.34 | 3.90 | 3.47 | 4.39 | 3.48 | 3.17 | 3.82 | 3.37 | 3.07 | 3.70 |
| Worries |  |  |  |  |  |  |  |  |  |  |  |  |  |  |  |
| Financial | 1.46 | 1.40 | 1.52 | 1.35 | 1.22 | 1.49 | 1.35 | 1.24 | 1.47 | 1.72 | 1.59 | 1.86 | 1.38 | 1.27 | 1.50 |
| Accessing essentials | 1.18 | 1.13 | 1.24 | 1.43 | 1.28 | 1.59 | 1.06 | 0.97 | 1.15 | 0.99 | 0.92 | 1.08 | 1.39 | 1.28 | 1.50 |
| COVID-19 illness | 0.94 | 0.90 | 0.98 | 0.82 | 0.74 | 0.90 | 0.80 | 0.74 | 0.87 | 0.97 | 0.90 | 1.05 | 1.12 | 1.04 | 1.22 |
| Social/ relationship concerns | 1.62 | 1.56 | 1.69 | 1.92 | 1.73 | 2.14 | 1.67 | 1.54 | 1.82 | 1.62 | 1.50 | 1.75 | 1.44 | 1.33 | 1.55 |
| Threats to safety | 1.42 | 1.35 | 1.48 | 1.70 | 1.52 | 1.90 | 1.41 | 1.28 | 1.54 | 1.42 | 1.31 | 1.54 | 1.31 | 1.20 | 1.42 |
| Number of observations | 206,714 |  |  | 16,495 |  |  | 53,752 |  |  | 74,635 |  |  | 61,832 |  |  |
| Number of individuals | 11,580 |  |  | 1,168 |  |  | 3,394 |  |  | 4,069 |  |  | 2,949 |  |  |

Note. Adversity experiences and worries variables are weighted to the proportions of gender, age, ethnicity, country, and education obtained from the Office for National Statistics. Individual adversity experiences and worries variables are binary. Analyses were further adjusted for day of the week and time since lockdown began. OR = odds ratio. CI = confidence interval.

**Table 4.** Fixed-effects logistic regression models predicting within-individual change in self-harm behaviours from individual categories of adversity experiences and worries

| Self-harm behaviours |  |  |  |  |  |  |  |  |  |  |  |  |  |  |  |
| --- | --- | --- | --- | --- | --- | --- | --- | --- | --- | --- | --- | --- | --- | --- | --- |
|  | Total sample |  |  | Ages 18-29 |  |  | Ages 30-44 |  |  | Ages 45-59 |  |  | Ages 60+ |  |  |
| Variable | OR | 95%<br>CI<br>lower | 95%<br>CI<br>upper | OR | 95%<br>CI<br>lower | 95%<br>CI<br>upper | OR | 95%<br>CI<br>lower | 95%<br>CI<br>upper | OR | 95%<br>CI<br>lower | 95%<br>CI<br>upper | OR | 95%<br>CI<br>lower | 95%<br>CI<br>upper |
| Adversity experiences |  |  |  |  |  |  |  |  |  |  |  |  |  |  |  |
| Financial | 1.01 | 0.93 | 1.11 | 1.01 | 0.85 | 1.21 | 0.80 | 0.66 | 0.97 | 1.10 | 0.95 | 1.27 | 1.14 | 0.93 | 1.41 |
| Accessing essentials | 1.18 | 1.06 | 1.31 | 1.10 | 0.89 | 1.36 | 0.88 | 0.70 | 1.12 | 1.36 | 1.14 | 1.64 | 1.43 | 1.14 | 1.79 |
| COVID-19 illness | 1.36 | 1.21 | 1.53 | 2.43 | 1.82 | 3.23 | 1.03 | 0.81 | 1.32 | 1.52 | 1.25 | 1.84 | 0.98 | 0.74 | 1.29 |
| Illness of others/<br>bereavement | 1.23 | 1.11 | 1.36 | 1.51 | 1.23 | 1.85 | 1.20 | 0.95 | 1.52 | 1.14 | 0.96 | 1.35 | 1.15 | 0.93 | 1.41 |
| Physical/ psychological<br>abuse | 4.94 | 4.57 | 5.35 | 3.39 | 2.84 | 4.06 | 6.96 | 5.84 | 8.30 | 4.34 | 3.78 | 4.96 | 5.96 | 5.09 | 6.98 |
| Worries |  |  |  |  |  |  |  |  |  |  |  |  |  |  |  |
| Financial | 1.22 | 1.13 | 1.32 | 1.11 | 0.96 | 1.29 | 1.28 | 1.09 | 1.50 | 1.40 | 1.21 | 1.60 | 1.12 | 0.94 | 1.32 |
| Accessing essentials | 1.22 | 1.14 | 1.31 | 1.47 | 1.27 | 1.70 | 1.09 | 0.93 | 1.27 | 1.17 | 1.02 | 1.34 | 1.18 | 1.01 | 1.37 |
| COVID-19 illness | 0.95 | 0.88 | 1.03 | 1.04 | 0.89 | 1.22 | 0.76 | 0.64 | 0.89 | 0.96 | 0.84 | 1.10 | 1.09 | 0.93 | 1.28 |
| Social/relationship<br>concerns | 1.31 | 1.21 | 1.42 | 1.79 | 1.52 | 2.10 | 1.14 | 0.97 | 1.34 | 1.33 | 1.16 | 1.53 | 1.11 | 0.93 | 1.32 |
| Threats to safety | 1.25 | 1.16 | 1.35 | 1.12 | 0.95 | 1.31 | 1.70 | 1.44 | 2.01 | 1.26 | 1.10 | 1.43 | 1.11 | 0.95 | 1.30 |
| Number of observations | 63,767 |  |  | 7,687 |  |  | 18,594 |  |  | 23,859 |  |  | 13,627 |  |  |
| Number of individuals | 3,747 |  |  | 530 |  |  | 1,201 |  |  | 1,329 |  |  | 687 |  |  |

Note. Adversity experiences and worries variables are weighted to the proportions of gender, age, ethnicity, country, and education obtained from the Office for National Statistics. Individual adversity experiences and worries variables are binary. Analyses were further adjusted for day of the week and time since lockdown began. OR = odds ratio. CI = confidence interval.

In the two younger age groups (18-29 and 30-44) and in older adults, social and relationship concerns had the second strongest associations with self-harm thoughts (OR range = 1.67 to 1.92) after experiencing physical/psychological abuse. In adults ages 45-59, the second strongest associations with self-harm thoughts after physical/psychological abuse were for financial concerns (OR = 1.72; 95% CI = 1.59 to 1.86).

In older adults (ages 60+), the second strongest association with self-harm behaviours was for having not been able to access essential items (OR = 1.43; 95% CI = 1.14 to 1.79), whilst this was the case for having had COVID-19 in adults ages 45-59 (OR = 1.52; 95% CI = 1.25 to 1.84) and young adults (OR = 2.43; 95% CI = 1.83 to 3.23), and threats to personal safety in the 30-44 age group (OR = 1.70; 95% CI = 1.44 to 2.01).

### Sensitivity analyses

When accounting for anxiety and depression symptoms within models, results were largely similar (Supplemental Tables S6-S9). Analyses examining physical abuse and psychological abuse as individual adversity experiences showed different patterns of association for each abuse type with outcomes (Supplemental Tables S10 and S11).

## Discussion

Both experiencing adversities and worrying about adversities were associated with an increased likelihood of self-harm thoughts and actually engaging in self-harm behaviours across the first 59 weeks of the COVID-19 pandemic. These results were found across age groups, with strongest associations for the total number of adversity experiences compared to the number of worries about adversities. The proportion in our sample reporting self-harm thoughts (26.1%) and self-harm behaviours (7.9%) at least once over the first year of the pandemic was higher than in other population-based studies conducted in the first few months of the pandemic, when approximately 10%^13,14^ of adults reported suicidal/self-harm thoughts and around 1%^13^ reported self-harm behaviours. Our findings are, however, similar to one US study which found that 31% of adults had reported thoughts of suicide/self-harm in the past two weeks.^18^

The largest predictor by far of both thinking about and engaging in self-harm was experiencing physical or psychological abuse, and this finding was consistent across all four age groups examined. Sensitivity analyses suggested that physical abuse is making larger contributions than psychological abuse to self-harm behaviours, whilst the size of the associations of both abuse types with self-harm thoughts were more similar. A range of literature outside of pandemic circumstances shows that different forms of abuse including domestic violence are predictors of self-harm behaviours and suicide.^26,27^ That increases in domestic abuse would occur during stay-at-home orders was anticipated early in the pandemic,^28^ and has been demonstrated in countries internationally. For example, the number of calls to emergency domestic abuse hotlines in the European Union had increased 60% by the end of April 2020,^29^ whilst intimate partner violence against women increased 23% over the first three months of the first lockdown in Spain.^30^

Worsening economic circumstances have been identified as one of the causes of this increase in domestic abuse,^30^ and financial stress was another predictor of self-harm thoughts and behaviours identified by our study and by other research conducted during the current pandemic.^18^ Notably, worrying about financial adversity such as losing one’s job rather than actually experiencing such an adversity was more consistently associated with self-harm thoughts and behaviours across age groups. This suggests that, thus far, economic uncertainty rather than actual adversities are negatively impacting people’s mental health, and it is possible that these associations may change in magnitude as the economic consequences of the pandemic and recession unfold. Considerable evidence indicates that economic recessions are associated with increases in rates of self-harm and suicide, particularly in the working-age population.^8^ Risk for both attempted suicide and suicide death is higher for those unemployed over the long-term compared to shorter term unemployment.^31,32^ However, an increase in self-harm and suicides during economic recessions is not inevitable.^1^

Lessons learnt from prior economic recessions suggest multiple opportunities for how governments can respond with policies to mitigate the mental health impact of the upcoming recession.^5^ The increases in suicides that correspond to unemployment rates are not uniform across all countries but are instead modified by differential investment in social programmes to mitigate these effects.^33^ For example, following the 2008 financial crisis, an increase of 1% per capita in government spending designed to mitigate the effect of financial hardship was associated with a 0.2% decrease in suicide in Japan.^34^ In the three decades leading up to the 2008 recession, every $10 USD invested per person on programmes aimed to increase chances of gainful employment resulted in a decrease of the effect of unemployment on suicides decreased by 0.04% in EU countries.^33^ Thus, our findings highlight the potential danger of the economic impacts of COVID-19 on self-harm thoughts and behaviours and, as self-harm is an important risk factor for suicide, potentially for suicide too, and suggest the importance of addressing economic concerns amongst individuals urgently.

Whilst many of our findings were consistent across age groups, there were some discrepancies. For example, worrying about catching COVID-19 was associated with reduced likelihood of having self-harm thoughts and self-harm behaviours in adults aged 30 to 44, but this pattern was reversed in the oldest age group (60 years plus), where worries about falling ill were associated with increased likelihood of thoughts of harming themselves or that they would be better off dead (self-harm thoughts). This could have been influenced by public health messaging highlighting that older adults are at higher risk than younger adults for dying of the illness; a death that news coverage often portrayed as occurring alone and without the ability to say goodbye to loved ones. In contrast, having already had the illness was related to increased likelihood of both outcomes in the total sample, and this remained true in young (18-29) and middle aged (45-59) adults for self-harm thoughts and for self-harm behaviours. It is therefore possible that the disease itself may play a role in increasing risk for self-harm thoughts and behaviours, while those who are particularly worried about falling ill with COVID-19 are more protective of their health and less likely to want to harm themselves. Evidence for the former, that COVID-19 illness leads to increased risk for mental health problems such as depression, self-harm thoughts and self-harm behaviours has been documented across a range of studies.^12,35^

This study has a number of strengths including the use of a large, well-stratified sample on socio-demographic groups which were weighted on the basis of population estimates of core demographics, and its longitudinal follow-up with repeated assessments of adversities, worries and self-harm thoughts and behaviours. We also used robust statistical methods to account for unobserved stable participant characteristics. However, sampling was not random and the data are therefore not representative of the general UK population. It is possible that individuals who were experiencing greater adversity and were more likely to have self-harm thoughts and behaviours were more likely to participate in the study. Nevertheless, this study did not aim to report prevalence of such experiences, but rather to identify the time-varying relationship between exposures and self-harm thoughts and behaviours. Crucially, the sample was heterogeneous and maintained its heterogeneity over time. There may have been other relevant forms of adversity and worry not captured in the current study, which may have resulted in an overestimation of the adversities and worries we did include. Finally, fixed effects regression does not address direction of causality. Arellano Bond models can be used to follow up fixed effects models to account for this lack of directionality,^36^ but because linear estimation is used, we could not utilise this approach as our outcomes were binary. However, whilst the relationship between worries and self-harm thoughts and behaviours may have involved some bidirectionality, there is little evidence to suggest that self-harm thoughts or behaviours increase the likelihood of individuals experiencing adversities.

Across the COVID-19 pandemic, there are concerns about potential future increases in suicide levels. Self-harm thoughts and behaviours are important and strong predictors of future suicide risk, so identifying modifiable risk factors for self-harm that can be addressed through public health interventions during the COVID-19 pandemic and beyond is vital.^1,5^ Our findings suggest that increases in self-harm thoughts and behaviours across the first 59 weeks of the pandemic were related to financial uncertainty, physical or psychological abuse, concern for others, not being able to access essential items, and worries about one’s personal safety. Suggestions have already been made for how to adapt evidence-based suicide prevention strategies to current the pandemic.^37^ For example, it has been recommended that universal interventions to mitigate the impact of poverty and unemployment on suicide risk should be implemented.^5^ Our data suggest the importance of following such strategies to try and reduce self-harm thoughts and behaviours that have the potential to drive increasing suicide rates of the coming months. The findings here also suggest the need for ongoing surveillance of how these well-established risk factors for suicide and self-harm may be exacerbated by the upcoming recession and as public health measures such as social distancing continue.

## Data Availability

The COVID-19 Social Study documentation and codebook are available for download at https://www.covidsocialstudy.org/. Statistical code is available upon request from Elise Paul.

## Acknowledgements

The researchers are grateful for the support of a number of organisations with their recruitment efforts including: the UKRI Mental Health Networks, Find Out Now, UCL BioResource, SEO Works, FieldworkHub, and Optimal Workshop.

## Author contribution

DF and EP conceptualised and designed the study. DF acquired funding, led the investigation, provided oversight on the methodology, provided software, and supervised the project. Data were curated, validated, and formally analysed by EP. EP created visualisations, wrote the original manuscript draft with input from all authors, who then reviewed and edited the manuscript. All authors approved the final version of the manuscript and had full access to and verified the data.

## Supplementary Materials

### Sampling methods

Three primary recruitment approaches were used. First, convenience sampling was used, including promoting the study through existing networks and mailing lists (including large databases of adults who had previously consented to be involved in health research across the UK), print and digital media coverage, and social media. Second, more targeted recruitment was undertaken focusing on groups who were anticipated to be less likely to take part in the research via our first strategy, including (i) individuals from a low-income background, (ii) individuals with no or few educational qualifications, and (iii) individuals who were unemployed. Third, the study was promoted via partnerships with third sector organisations to vulnerable groups, including adults with pre-existing mental health conditions, older adults, carers, and people experiencing domestic violence or abuse. Recruitment was refreshed in August when participants who were lost-to-follow-up were recontacted. The study was approved by the UCL Research Ethics Committee [12467/005] and all participants gave informed consent. Participants were not compensated for participation.

### Outcomes

#### Self-harm thoughts and behaviours

Two outcome variables were assessed weekly for the first 22 weeks of the study (1 April to 21 August 2020) and then monthly to 17 May 2021. Thoughts of death or self-harm (“self-harm thoughts”) were measured with an item from the Patient Health Questionnaire (PHQ-9)^1^, an instrument often used as a screening tool for depression in primary care practice: “Over the last week, how often have you been bothered by: Thoughts that you would be better off dead or hurting yourself in some way?”. Second, self-harm behaviours were measured with a similar study-developed item: “Over the last week, how often have you been bothered by: Self-harming or deliberately hurting yourself?”. Responses to both items were rated on a four-point scale from “not at all” to “nearly every day”. Analyses focused on binary variables indicating the presence of at least some self-harm thoughts and self-harm behaviours at each time point.

### Exposure variables

#### Adversities

Financial adversity consisted of: (i) loss of job/been unable to do paid work; (ii) spouse/partner lost their job/was unable to do paid work; (iii) major cut in household income (e.g., due to you or your partner being furloughed/put on leave/ not receiving sufficient work); (iv) unable to pay bills/rent/mortgage; and (v) evicted/lost accommodation. Illness with COVID-19 was measured with an item asking if participants had either suspected or been diagnosed with the illness. Family/friend illness or bereavement was assessed as: (i) the participant having someone close to them ill in hospital (due to COVID-19 or another cause), and (ii) lost somebody close (due to COVID-19 or another cause). Participants indicated whether they had been “physically harmed or hurt by someone else” or “bullied, controlled, intimidated, or psychologically hurt by someone else” during the last week. Responses were rated on a four-point scale ranging from “not at all” to “nearly every day”. A response to either item indicating physical or psychological abuse was categorised as present. Finally, inability to access essential items was measured by asking if participants had been unable to access sufficient food and required medication. Total weekly adversities scores were calculated by summing each of the five binary variables (range 0-5).

#### Worries about adversity

Two questions asked participants to select which of a list of items had caused them (i) stress (however minor) in the past week, and (ii) significant stress (something being constantly on their mind or keeping them awake at night in the past week). Responses were classified as worries whether participants said they were causing them either minor or significant stress. Financial worries was measured with three items: (i) losing your job/unemployment, (ii) finances, and (iii) work, COVID-19 illness with two: (i) catching COVID-19 and (ii) becoming seriously ill from COVID-19, social and relationship worries with five: (i) marriage or other romantic relationship, (ii) friends or family living in your household, (iii) friends or family living outside your household, (iv) neighbours, and (v) your pet), safety and security concerns with one: your own safety/security’, and worries about access essentials with two (i) food and (ii) medication. Total weekly worries scores were computed by summing these five binary variables (range 0-5).

#### Variables used to describe the samples

Several socio-demographic and health factors were used to describe the two samples. All were measured when participants first joined the study. These included gender (women vs men), ethnicity (white vs ethnic minority groups), age groups (age 18-29, 30-45, 46-59, 60+) and education (up to GCSE levels, A-levels or equivalent, and university degree or above). For ethnicity, participants were asked: “What is your ethnicity?”. Answer choices were: i) Asian/Asian British - Indian, Pakistani, Bangladeshi, other, ii) Black/Black British - Caribbean, African, other, iii) Mixed race - White and Black/Black British, iv) Mixed race – other, v) White - British, Irish, other, vi) Chinese/Chinese British, vii) Middle Eastern/Middle Eastern British - Arab, Turkish, other, viii) Other ethnic group, and ix) Prefer not to say. Respondents endorsing v (White- British, Irish, other) were classified as white, whilst all other categories were classified as ethnic minority groups.

We also included two health-related factors to describe the samples: self-reported diagnosis of any long-term physical health condition (e.g., asthma or diabetes) or any disability (yes vs no), and self-reported diagnosis of any long-term mental health condition (e.g., depression, anxiety) (yes vs no). Participants were asked if they had at least one of eight long-term physical health conditions (high blood pressure, diabetes, heart disease, lung disease [e.g., asthma or COPD], cancer, another clinically-diagnosed chronic physical health condition). They were also asked if they had at least one of four long-term mental health conditions (clinically-diagnosed depression, clinically-diagnosed anxiety, another clinically-diagnosed mental health problem).

### Statistical Analysis

The basic model can be expressed as follows:

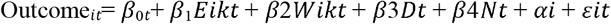

where Outcome_*it*_ is a measure of individual *i*’s self-harm thoughts or self-harm behaviours at time *t*, E is whether an individual *i* was experiencing adversity at time, W is whether an individual was worrying about adversity *k* at time *t*, D*t* is a vector of indicator variables for day, *Nt* is a continuous variable for days since lockdown, is unobserved time invariant confounding factors, and is *ε* error.

Three sets of sensitivity analyses were conducted to ensure the robustness of our results. First, models with individual categories of adversities and worries were re-run which included continuous measures of weekly depression symptoms assessed with the PHQ-9^1^, without the inclusion of the self-harm thoughts item. Second, these models were estimated again, but this time with continuous measures of weekly anxiety symptoms using the Generalized Anxiety Disorder (GAD-7) scale^2^. Third, we repeated our fixed effects models using individual categories of worry and adversity with the physical/psychological abuse variable separated into two separate variables: physical abuse and psychological abuse.

To account for the non-random nature of the sample and increase representativeness of the UK general population, all data were weighted to the proportions of gender, age, ethnicity, country, and education obtained from the Office for National Statistics^3^. Weights were constructed using a multivariate reweighting method using the Stata user written command ‘ebalance’^4^. Analyses were conducted using Stata version 16^5^ and fixed effects model results plotted with the user-written command ‘coefplot’^6^. Full details on sampling, recruitment, data collection, data cleaning, weighting and sample demographics are available at https://github.com/UCL-BSH/CSSUserGuide.

## Results

### Sensitivity analyses

When accounting for anxiety and depression symptoms within models, results were largely similar. But financial adversities were no longer increased risk for either outcome in any age group and were related to decreased likelihood of outcomes in some age groups (Supplemental Tables S6-S9). Worries about financial adversity, however, remained associated with increased likelihood of both outcomes in the total sample and across some age groups. Analyses examining physical abuse and psychological abuse as individual adversity experiences showed different patterns of association for each abuse type with outcomes (Supplemental Tables S10 and S11). Whilst physical abuse (OR range = 5.77 to 19.53) had much larger associations than psychological abuse (OR range = 1.74 to 2.69) with self-harm behaviours in all age groups, the associations for these two abuse types were more similar in magnitude across age groups for self-harm thoughts.

**Figure S1a.**
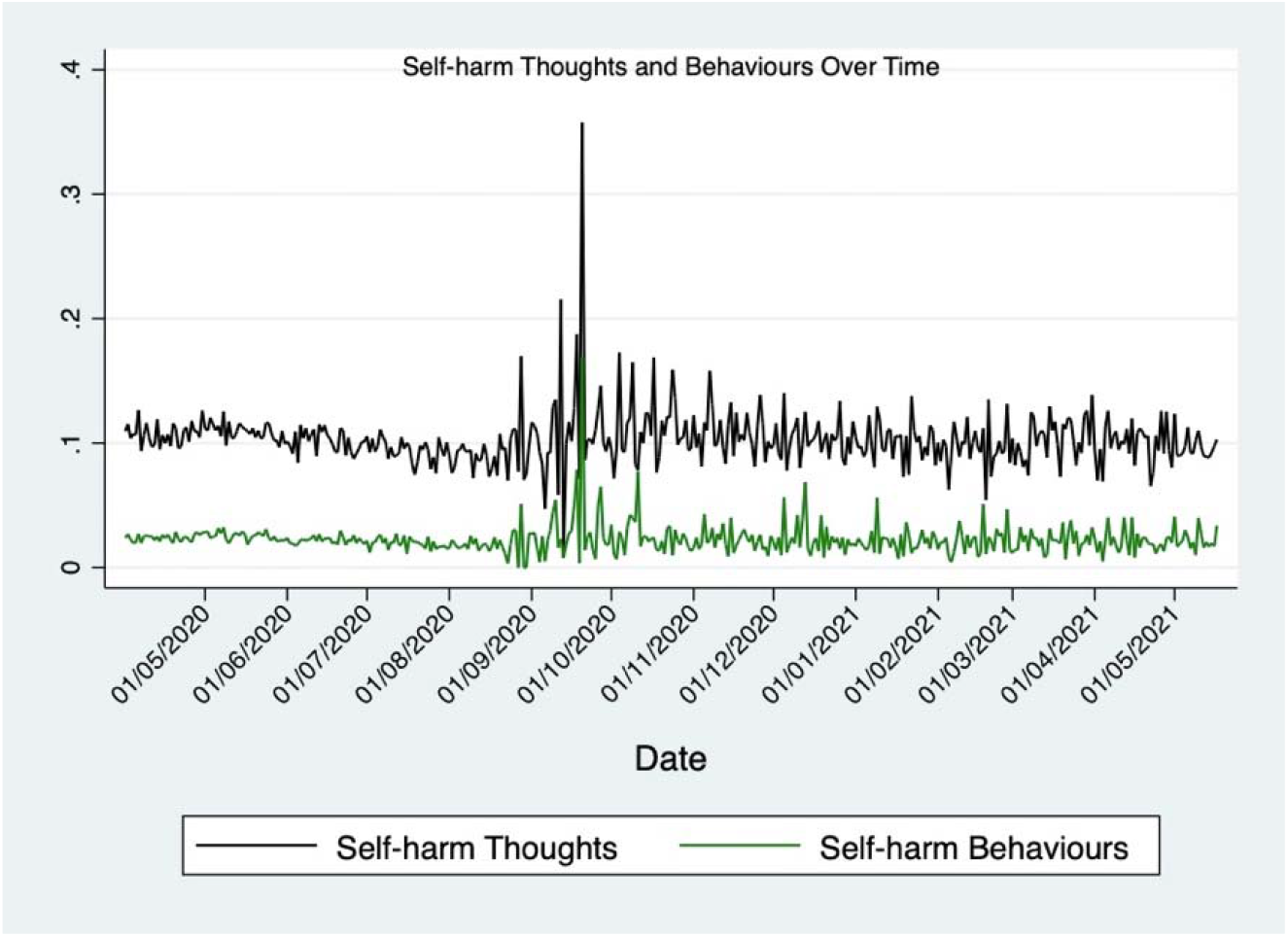
Average proportion of the sample with self-harm thoughts and self-harm behaviours over time for the duration of the study period (1 April 2020 to 17 May 2021). Increased variability in data starting in August may be due to the change in data collection frequency from weekly to monthly that occurred at this time.

**Figure S1b.**
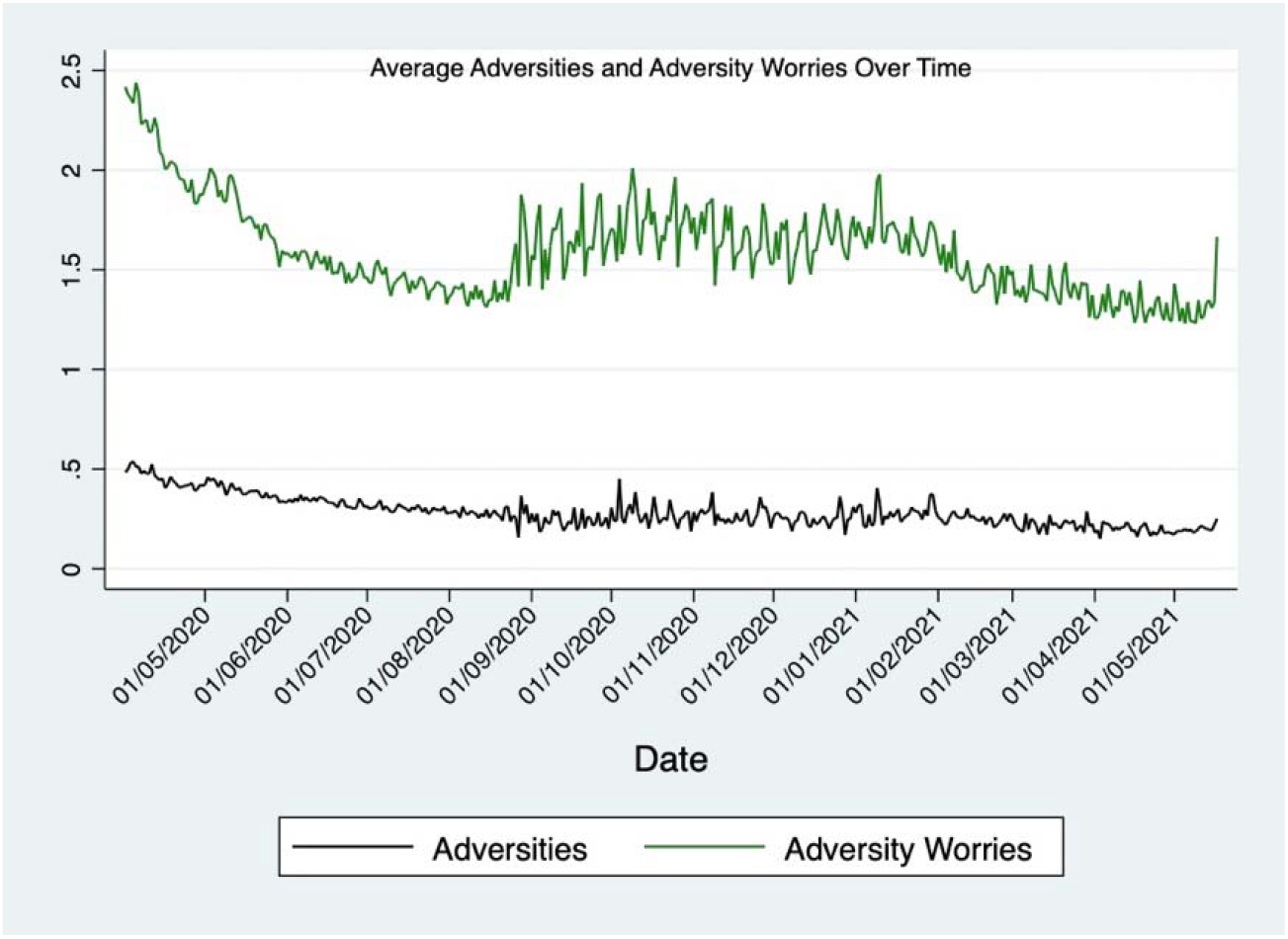
Mean adversities and worries about adversity over time for the duration of the study period (1 April 2020 to 17 May 2021). Increased variability in data starting in August may be due to the change in data collection frequency from weekly to monthly that occurred at this time.

**Figure S2.**
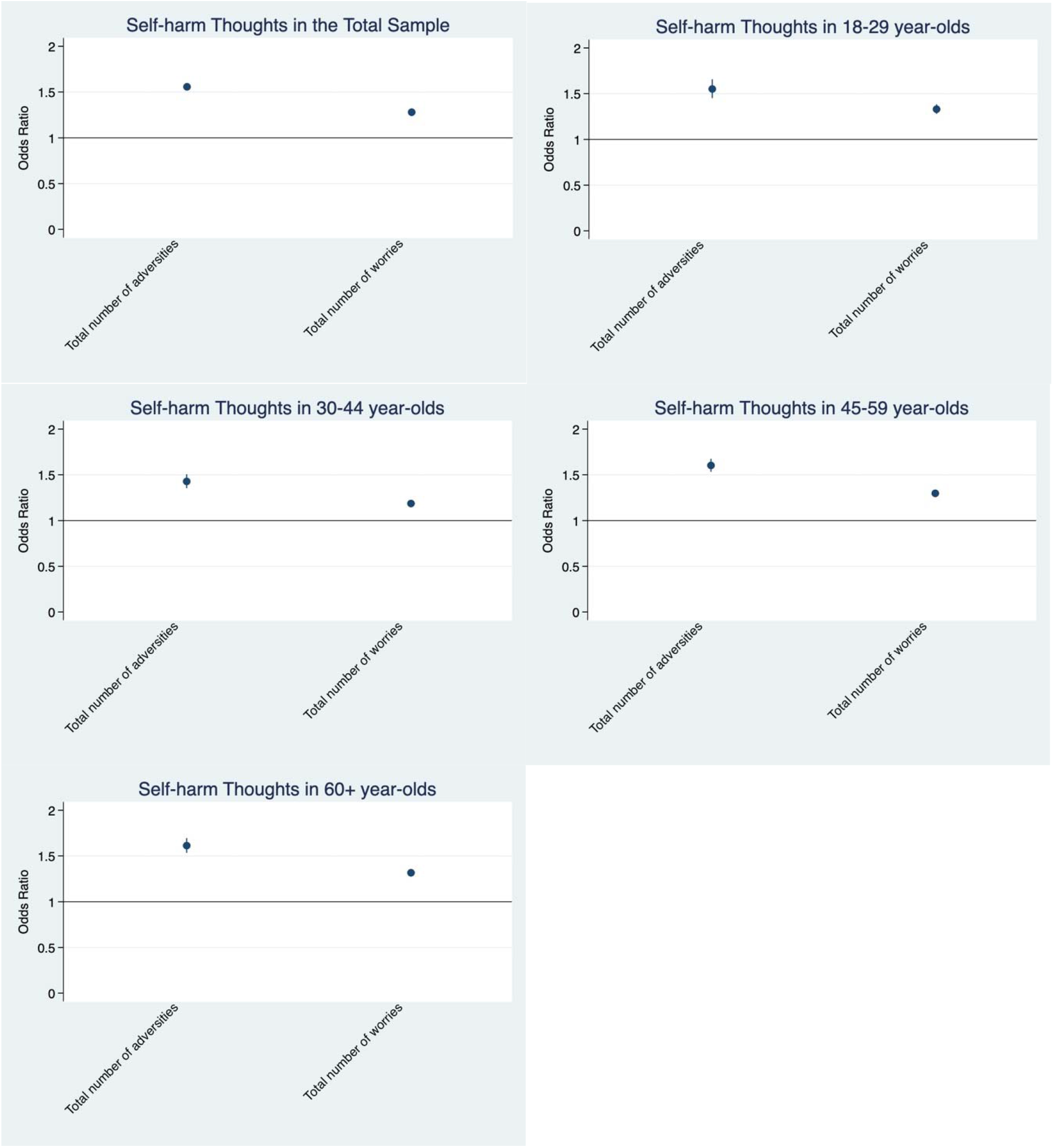
Associations between the total number adversity experiences and worries about adversity and change over time in self-harm thoughts derived from fixed effects models. Experiences and worries were entered simultaneously into the same model. Analyses were further adjusted for day of the week and time since lockdown began.

**Figure S3.**
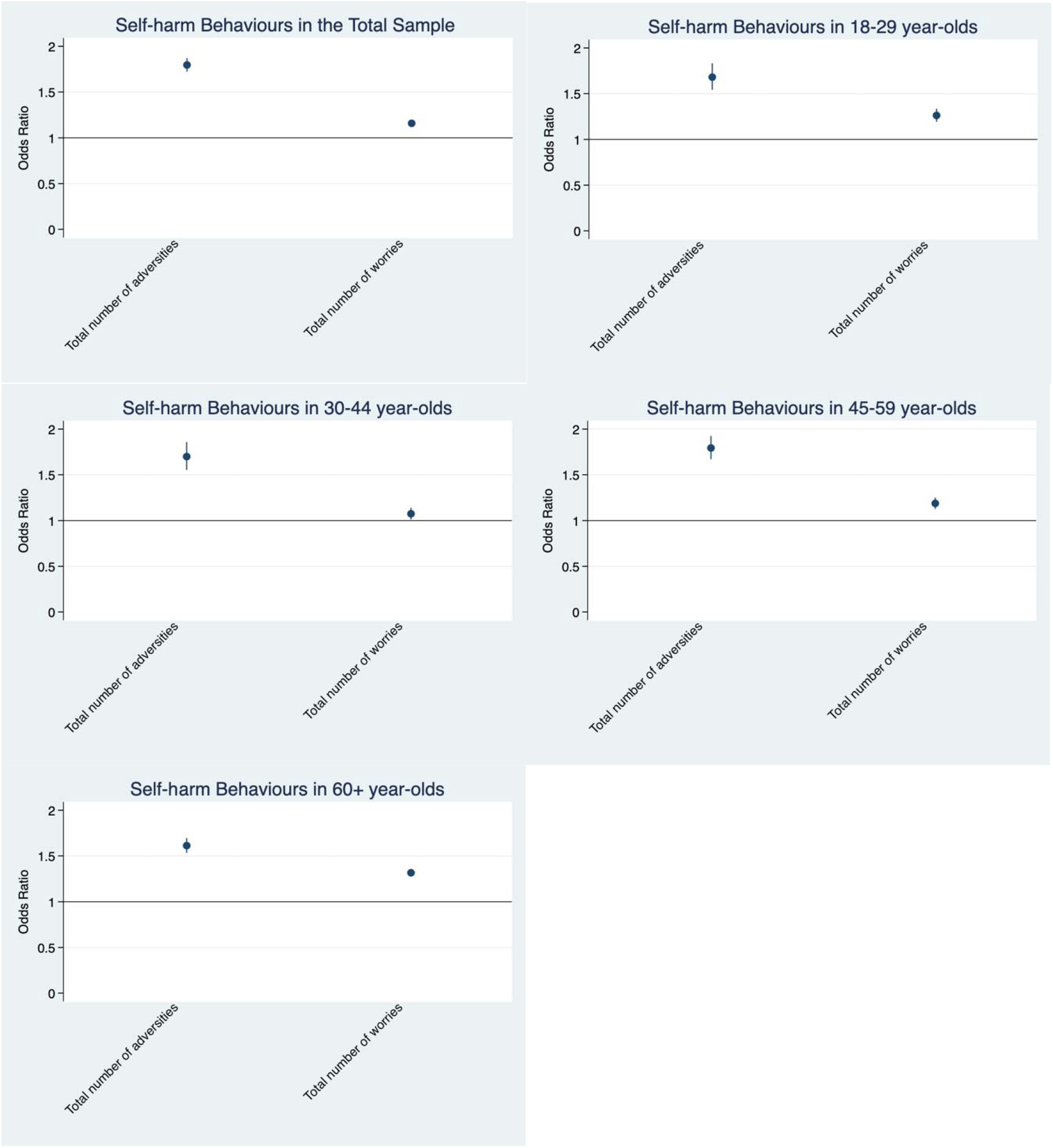
Associations between the total number adversity experiences and worries about adversity and change over time in self-harm behaviours derived from fixed effects models stratified by age. Experiences and worries were entered simultaneously into the same model. Analyses were further adjusted for day of the week and time since lockdown began.

**Figure S4.**
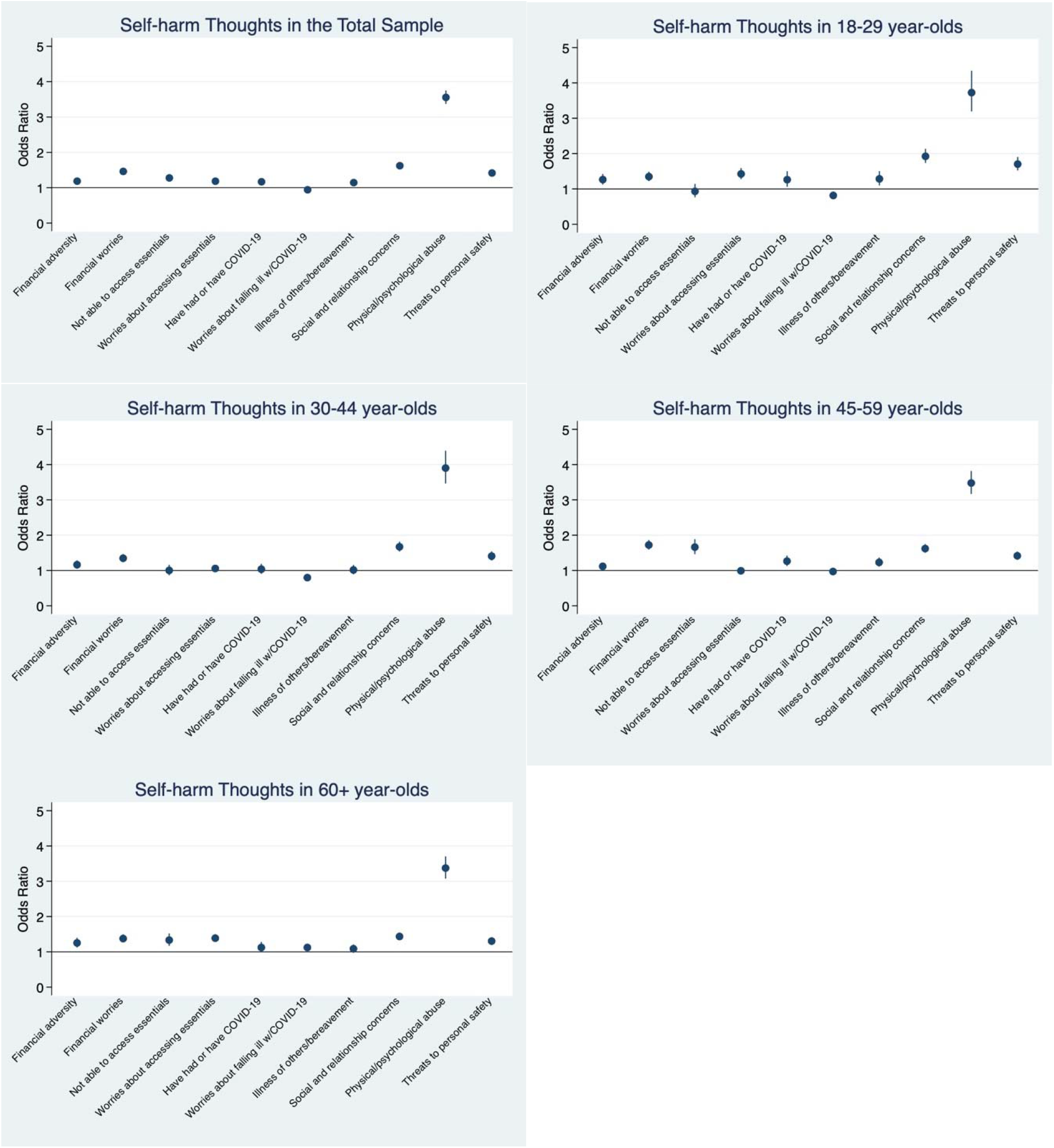
Associations between individual adversity experiences and worries about adversity and change over time in self-harm thoughts derived from fixed effects models. Experiences and worries were entered simultaneously into the same model. Analyses were further adjusted for day of the week and time since lockdown began.

**Figure S5.**
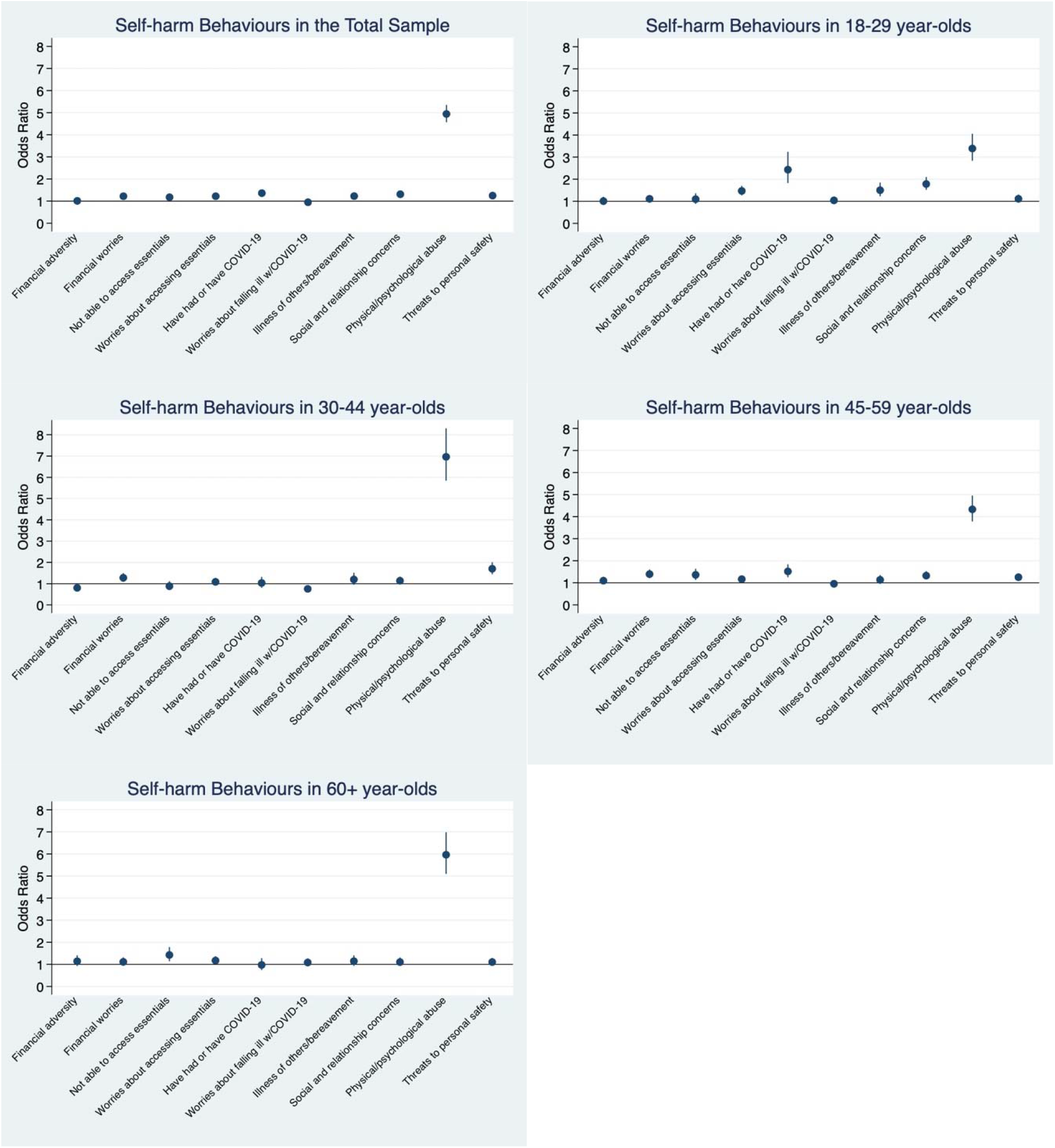
Associations between individual adversity experiences and worries about adversity and change over time in self-harm behaviours derived from fixed effects models. Experiences and worries were entered simultaneously into the same model. Analyses were further adjusted for day of the week and time since lockdown began.

**Table S1.**
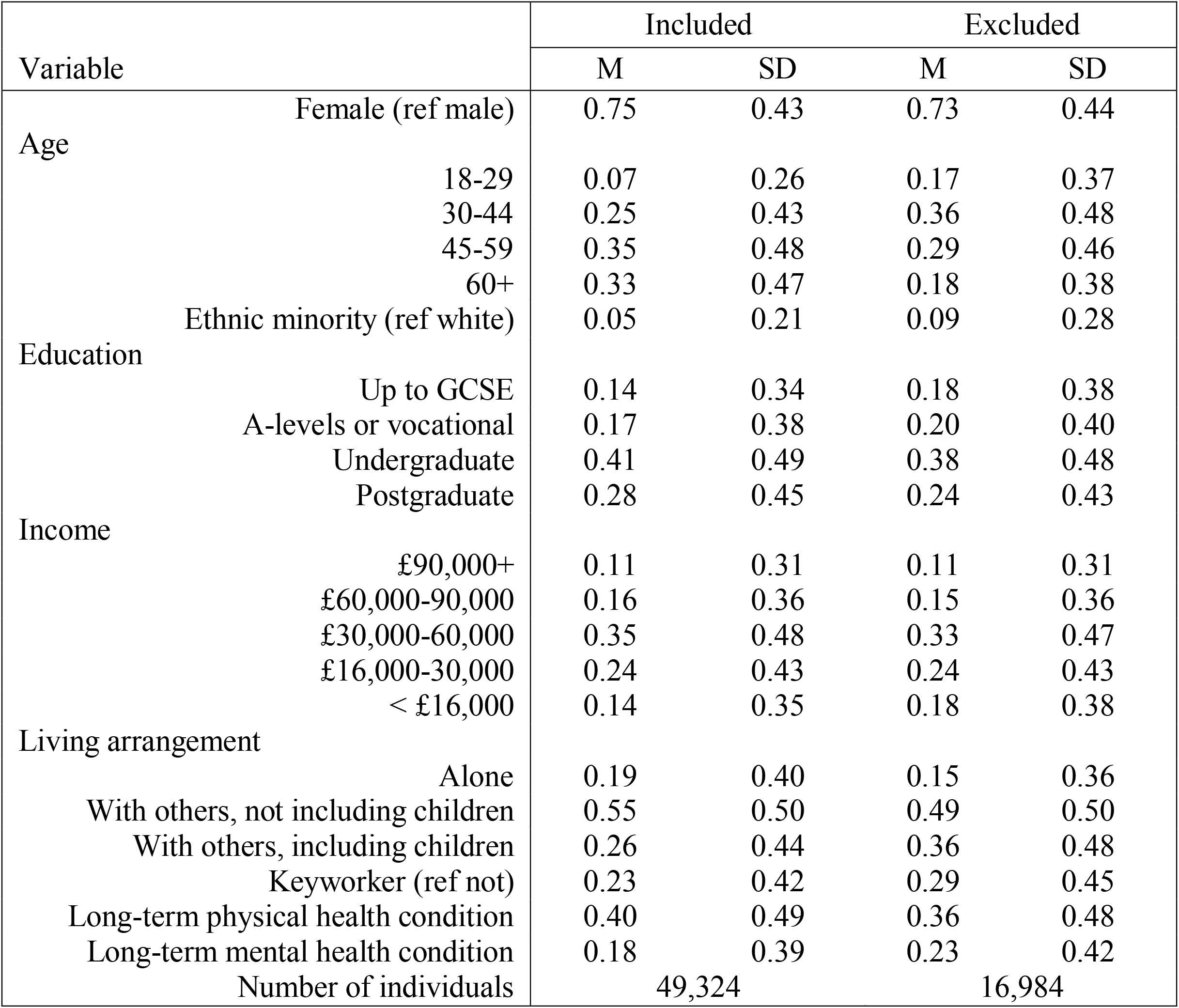
Characteristics of included and excluded participants, unweighted

**Table S2.**
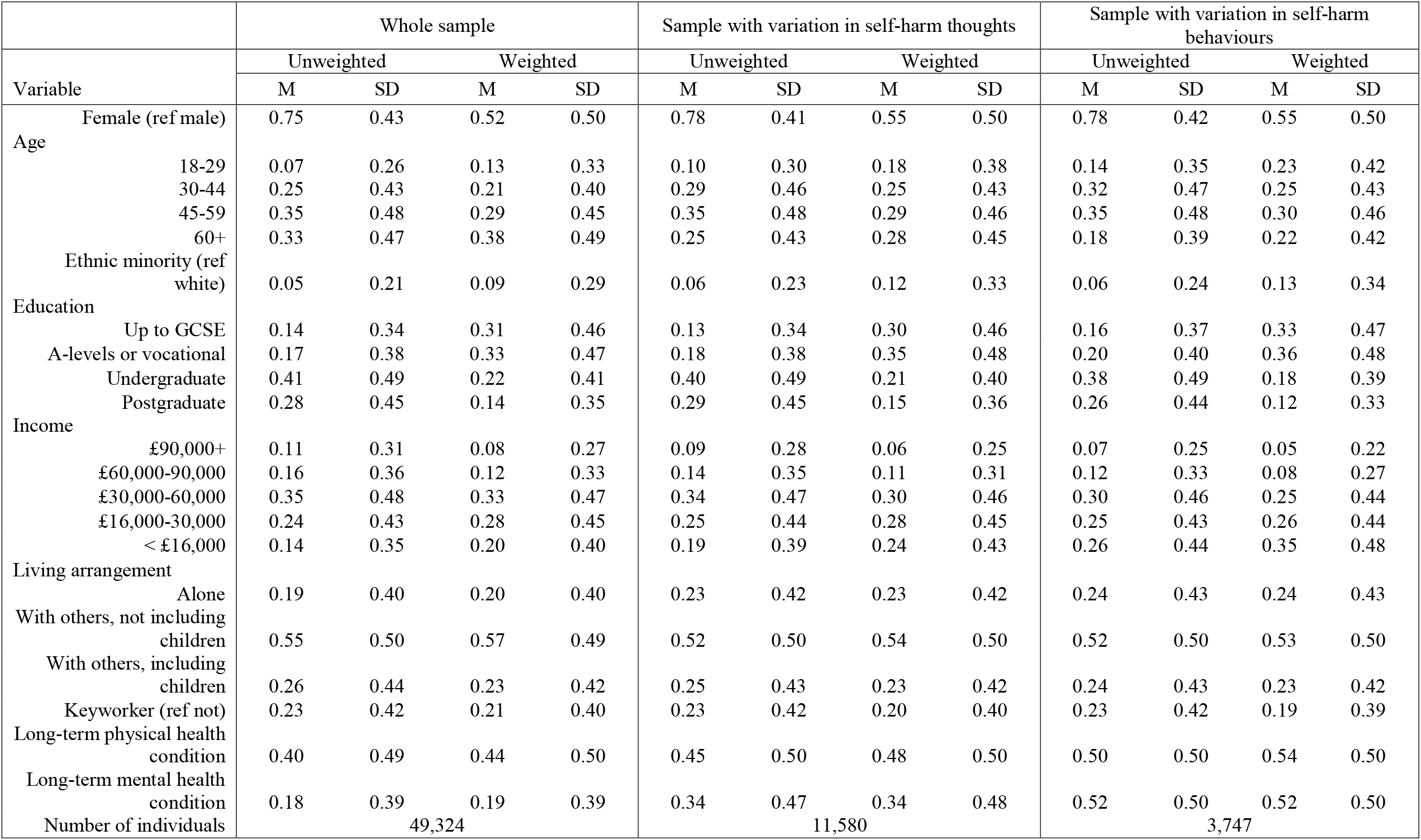
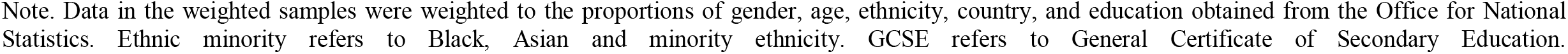
Socio-demographic characteristics for entire sample and for those with variation in self-harm thoughts and self-harm behaviours, unweighted and weighted

|  | Whole sample |  |  |  | Sample with variation in self-harm thoughts |  |  |  | Sample with variation in self-harm behaviours |  |  |  |
| --- | --- | --- | --- | --- | --- | --- | --- | --- | --- | --- | --- | --- |
| Variable | Unweighted |  | Weighted |  | Unweighted |  | Weighted |  | Unweighted |  | Weighted |  |
|  | M | SD | M | SD | M | SD | M | SD | M | SD | M | SD |
| Female (ref male) | 0.75 | 0.43 | 0.52 | 0.50 | 0.78 | 0.41 | 0.55 | 0.50 | 0.78 | 0.42 | 0.55 | 0.50 |
| Age |  |  |  |  |  |  |  |  |  |  |  |  |
| 18-29 | 0.07 | 0.26 | 0.13 | 0.33 | 0.10 | 0.30 | 0.18 | 0.38 | 0.14 | 0.35 | 0.23 | 0.42 |
| 30-44 | 0.25 | 0.43 | 0.21 | 0.40 | 0.29 | 0.46 | 0.25 | 0.43 | 0.32 | 0.47 | 0.25 | 0.43 |
| 45-59 | 0.35 | 0.48 | 0.29 | 0.45 | 0.35 | 0.48 | 0.29 | 0.46 | 0.35 | 0.48 | 0.30 | 0.46 |
| 60+ | 0.33 | 0.47 | 0.38 | 0.49 | 0.25 | 0.43 | 0.28 | 0.45 | 0.18 | 0.39 | 0.22 | 0.42 |
| Ethnic minority (ref white) | 0.05 | 0.21 | 0.09 | 0.29 | 0.06 | 0.23 | 0.12 | 0.33 | 0.06 | 0.24 | 0.13 | 0.34 |
| Education |  |  |  |  |  |  |  |  |  |  |  |  |
| Up to GCSE | 0.14 | 0.34 | 0.31 | 0.46 | 0.13 | 0.34 | 0.30 | 0.46 | 0.16 | 0.37 | 0.33 | 0.47 |
| A-levels or vocational | 0.17 | 0.38 | 0.33 | 0.47 | 0.18 | 0.38 | 0.35 | 0.48 | 0.20 | 0.40 | 0.36 | 0.48 |
| Undergraduate | 0.41 | 0.49 | 0.22 | 0.41 | 0.40 | 0.49 | 0.21 | 0.40 | 0.38 | 0.49 | 0.18 | 0.39 |
| Postgraduate | 0.28 | 0.45 | 0.14 | 0.35 | 0.29 | 0.45 | 0.15 | 0.36 | 0.26 | 0.44 | 0.12 | 0.33 |
| Income |  |  |  |  |  |  |  |  |  |  |  |  |
| £90,000+ | 0.11 | 0.31 | 0.08 | 0.27 | 0.09 | 0.28 | 0.06 | 0.25 | 0.07 | 0.25 | 0.05 | 0.22 |
| £60,000-90,000 | 0.16 | 0.36 | 0.12 | 0.33 | 0.14 | 0.35 | 0.11 | 0.31 | 0.12 | 0.33 | 0.08 | 0.27 |
| £30,000-60,000 | 0.35 | 0.48 | 0.33 | 0.47 | 0.34 | 0.47 | 0.30 | 0.46 | 0.30 | 0.46 | 0.25 | 0.44 |
| £16,000-30,000 | 0.24 | 0.43 | 0.28 | 0.45 | 0.25 | 0.44 | 0.28 | 0.45 | 0.25 | 0.43 | 0.26 | 0.44 |
| < £16,000 | 0.14 | 0.35 | 0.20 | 0.40 | 0.19 | 0.39 | 0.24 | 0.43 | 0.26 | 0.44 | 0.35 | 0.48 |
| Living arrangement |  |  |  |  |  |  |  |  |  |  |  |  |
| Alone | 0.19 | 0.40 | 0.20 | 0.40 | 0.23 | 0.42 | 0.23 | 0.42 | 0.24 | 0.43 | 0.24 | 0.43 |
| With others, not including children | 0.55 | 0.50 | 0.57 | 0.49 | 0.52 | 0.50 | 0.54 | 0.50 | 0.52 | 0.50 | 0.53 | 0.50 |
| With others, including children | 0.26 | 0.44 | 0.23 | 0.42 | 0.25 | 0.43 | 0.23 | 0.42 | 0.24 | 0.43 | 0.23 | 0.42 |
| Keyworker (ref not) | 0.23 | 0.42 | 0.21 | 0.40 | 0.23 | 0.42 | 0.20 | 0.40 | 0.23 | 0.42 | 0.19 | 0.39 |
| Long-term physical health condition | 0.40 | 0.49 | 0.44 | 0.50 | 0.45 | 0.50 | 0.48 | 0.50 | 0.50 | 0.50 | 0.54 | 0.50 |
| Long-term mental health condition | 0.18 | 0.39 | 0.19 | 0.39 | 0.34 | 0.47 | 0.34 | 0.48 | 0.52 | 0.50 | 0.52 | 0.50 |
| Number of individuals | 49,324 |  |  |  | 11,580 |  |  |  | 3,747 |  |  |  |

Note. Data in the weighted samples were weighted to the proportions of gender, age, ethnicity, country, and education obtained from the Office for National Statistics. Ethnic minority refers to Black, Asian and minority ethnicity. GCSE refers to General Certificate of Secondary Education.

**Table S3.** Descriptive statistics by number of times self-harm thoughts were reported in total sample (N = 49,324), weighted

|  |  | Self-harm thoughts |  |  |  |  |  |
| --- | --- | --- | --- | --- | --- | --- | --- |
|  |  | Never<br>n = 36,455<br>(73.9%) |  | Once or twice<br>n = 5,650<br>(11.5%) |  | Thrice or more<br>n = 7,219<br>(14.6%) |  |
| Variable |  | M | SD | M | SD | M | SD |
| Gender (ref male) | Female | 0.51 | 0.50 | 0.55 | 0.50 | 0.53 | 0.50 |
| Age | 18-29 | 0.10 | 0.30 | 0.18 | 0.38 | 0.20 | 0.40 |
|  | 30-44 | 0.19 | 0.39 | 0.26 | 0.44 | 0.24 | 0.43 |
|  | 45-59 | 0.28 | 0.45 | 0.28 | 0.45 | 0.31 | 0.46 |
|  | 60+ | 0.43 | 0.49 | 0.27 | 0.44 | 0.25 | 0.43 |
| Ethnicity (ref white) | Ethnic minority | 0.08 | 0.28 | 0.13 | 0.33 | 0.12 | 0.32 |
| Education | Up to GCSE | 0.31 | 0.46 | 0.29 | 0.46 | 0.32 | 0.47 |
|  | A-levels or vocational | 0.32 | 0.47 | 0.35 | 0.48 | 0.36 | 0.48 |
|  | Undergraduate | 0.22 | 0.42 | 0.21 | 0.40 | 0.19 | 0.39 |
|  | Postgraduate | 0.15 | 0.35 | 0.15 | 0.36 | 0.13 | 0.34 |
| Income | £90,000+ | 0.09 | 0.27 | 0.08 | 0.26 | 0.05 | 0.22 |
|  | £60,000-90,000 | 0.13 | 0.33 | 0.12 | 0.33 | 0.08 | 0.27 |
|  | £30,000-60,000 | 0.34 | 0.47 | 0.30 | 0.46 | 0.28 | 0.45 |
|  | £16,000-30,000 | 0.28 | 0.45 | 0.28 | 0.45 | 0.28 | 0.45 |
|  | < £16,000 | 0.17 | 0.37 | 0.22 | 0.42 | 0.31 | 0.46 |
| Living arrangement | Alone | 0.18 | 0.38 | 0.20 | 0.40 | 0.26 | 0.44 |
|  | With others, not children | 0.59 | 0.49 | 0.54 | 0.50 | 0.54 | 0.50 |
|  | With others, including children | 0.23 | 0.42 | 0.26 | 0.44 | 0.21 | 0.40 |
| Keyworker (ref not) | Keyworker | 0.21 | 0.41 | 0.21 | 0.41 | 0.18 | 0.39 |
| Long-term physical health condition |  |  |  |  |  |  |  |
|  | Present | 0.42 | 0.49 | 0.45 | 0.50 | 0.52 | 0.50 |
| Long-term mental health condition |  |  |  |  |  |  |  |
|  | Present | 0.11 | 0.31 | 0.28 | 0.45 | 0.48 | 0.50 |

Note. Data were weighted to the proportions of gender, age, ethnicity, country, and education obtained from the Office for National Statistics. Ethnic minority refers to Black, Asian and minority ethnicity. GCSE refers to General Certificate of Secondary Education.

**Table S4.**
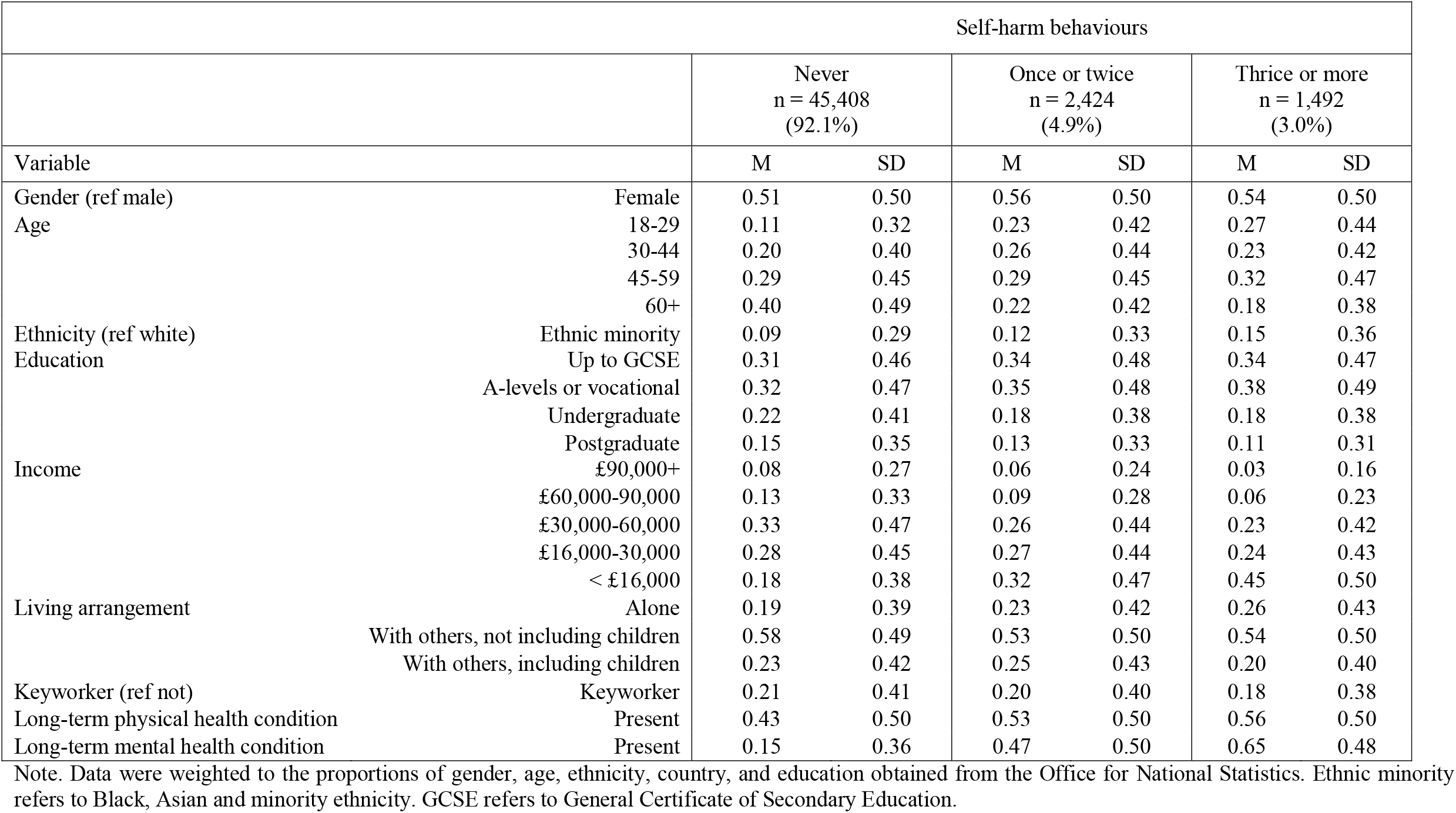
Descriptive statistics by number of times self-harm behaviours were reported in total sample (N = 49,324), weighted

**Table S5.** Descriptive statistics for predictor and outcome variables amongst individuals with variation in each outcome variable

|  |  | Sample with variation in self-harm thoughts |  |  |  | Sample with variation in self-harm behaviours |  |  |  |
| --- | --- | --- | --- | --- | --- | --- | --- | --- | --- |
| Variable |  | Overall Mean | Overall SD | Between SD | Within SD | Overall Mean | Overall SD | Between SD | Within SD |
| Outcomes | Self-harm thoughts | 0.28 | 0.45 | 0.26 | 0.36 | 0.47 | 0.50 | 0.36 | 0.34 |
|  | Self-harm behaviours | 0.04 | 0.20 | 0.14 | 0.16 | 0.20 | 0.40 | 0.23 | 0.33 |
| Adversity experiences |  |  |  |  |  |  |  |  |  |
|  | Total adversities (0-5) | 0.48 | 0.71 | 0.57 | 0.47 | 0.61 | 0.80 | 0.66 | 0.52 |
|  | Financial | 0.15 | 0.36 | 0.29 | 0.24 | 0.17 | 0.38 | 0.30 | 0.25 |
|  | Accessing essentials | 0.04 | 0.20 | 0.15 | 0.16 | 0.07 | 0.26 | 0.19 | 0.20 |
|  | COVID-19 illness | 0.12 | 0.32 | 0.27 | 0.19 | 0.12 | 0.33 | 0.27 | 0.20 |
|  | Illness of others/bereavement | 0.06 | 0.24 | 0.12 | 0.21 | 0.07 | 0.26 | 0.15 | 0.22 |
|  | Physical/psychological abuse | 0.11 | 0.31 | 0.23 | 0.22 | 0.17 | 0.38 | 0.28 | 0.26 |
| Worries |  |  |  |  |  |  |  |  |  |
|  | Total worries (0-5) | 2.18 | 1.21 | 0.96 | 0.78 | 2.36 | 1.28 | 1.01 | 0.81 |
|  | Financial | 0.60 | 0.49 | 0.37 | 0.31 | 0.64 | 0.48 | 0.36 | 0.31 |
|  | Accessing essentials | 0.19 | 0.39 | 0.27 | 0.30 | 0.25 | 0.43 | 0.30 | 0.33 |
|  | COVID-19 illness | 0.58 | 0.50 | 0.38 | 0.32 | 0.48 | 0.50 | 0.38 | 0.32 |
|  | Social/relationship concerns | 0.77 | 0.42 | 0.27 | 0.32 | 0.79 | 0.41 | 0.27 | 0.31 |
|  | Threats to safety | 0.14 | 0.35 | 0.24 | 0.27 | 0.20 | 0.40 | 0.28 | 0.30 |
| Number of observations |  | 206,714 |  |  |  | 63,767 |  |  |  |
| Number of individuals |  | 11,580 |  |  |  | 3,747 |  |  |  |

**Table S6.** Fixed-effects logistic regression models predicting within-individual change in self-harm thoughts from individual categories of adversity experiences and worries accounting for anxiety symptoms

| Self-harm thoughts |  |  |  |  |  |  |  |  |  |  |  |  |  |  |  |
| --- | --- | --- | --- | --- | --- | --- | --- | --- | --- | --- | --- | --- | --- | --- | --- |
|  | Total sample |  |  | Ages 18-29 |  |  | Ages 30-44 |  |  | Ages 45-59 |  |  | Ages 60+ |  |  |
| Variable | OR | 95%<br>CI<br>lower | 95%<br>CI<br>upper | OR | 95%<br>CI<br>lower | 95%<br>CI<br>upper | OR | 95%<br>CI<br>lower | 95%<br>CI<br>upper | OR | 95%<br>CI<br>lower | 95%<br>CI<br>upper | OR | 95%<br>CI<br>lower | 95%<br>CI<br>upper |
| Anxiety symptoms | 1.34 | 1.33 | 1.34 | 1.32 | 1.30 | 1.33 | 1.32 | 1.30 | 1.33 | 1.34 | 1.33 | 1.35 | 1.36 | 1.35 | 1.38 |
| Adversity experiences |  |  |  |  |  |  |  |  |  |  |  |  |  |  |  |
| Financial | 1.01 | 0.96 | 1.07 | 1.06 | 0.93 | 1.20 | 1.04 | 0.93 | 1.16 | 0.97 | 0.88 | 1.06 | 1.03 | 0.92 | 1.16 |
| Accessing essentials | 1.17 | 1.09 | 1.27 | 0.84 | 0.67 | 1.04 | 0.91 | 0.78 | 1.07 | 1.49 | 1.30 | 1.71 | 1.27 | 1.11 | 1.47 |
| COVID-19 illness | 1.11 | 1.03 | 1.20 | 1.19 | 0.99 | 1.44 | 1.00 | 0.86 | 1.15 | 1.23 | 1.09 | 1.40 | 1.05 | 0.91 | 1.22 |
| Illness of others/<br>bereavement | 0.97 | 0.91 | 1.04 | 1.07 | 0.90 | 1.26 | 0.92 | 0.80 | 1.06 | 1.06 | 0.95 | 1.19 | 0.89 | 0.79 | 1.00 |
| Physical/psychological<br>abuse | 2.68 | 2.52 | 2.83 | 3.28 | 2.78 | 3.87 | 3.01 | 2.65 | 3.42 | 2.59 | 2.35 | 2.87 | 2.36 | 2.13 | 2.61 |
| Worries |  |  |  |  |  |  |  |  |  |  |  |  |  |  |  |
| Financial | 1.16 | 1.11 | 1.22 | 1.11 | 1.00 | 1.23 | 1.05 | 0.96 | 1.16 | 1.30 | 1.19 | 1.41 | 1.18 | 1.08 | 1.29 |
| Accessing essentials | 1.02 | 0.97 | 1.07 | 1.20 | 1.06 | 1.35 | 0.92 | 0.84 | 1.01 | 0.86 | 0.79 | 0.94 | 1.18 | 1.09 | 1.29 |
| COVID-19 illness | 0.84 | 0.80 | 0.88 | 0.75 | 0.67 | 0.83 | 0.74 | 0.68 | 0.81 | 0.85 | 0.78 | 0.92 | 0.98 | 0.90 | 1.07 |
| Social/relationship<br>concerns | 1.31 | 1.26 | 1.38 | 1.53 | 1.36 | 1.71 | 1.34 | 1.22 | 1.47 | 1.30 | 1.20 | 1.41 | 1.22 | 1.12 | 1.33 |
| Threats to safety | 1.12 | 1.06 | 1.17 | 1.39 | 1.23 | 1.57 | 1.07 | 0.97 | 1.18 | 1.12 | 1.03 | 1.22 | 1.05 | 0.96 | 1.15 |
| Number of observations | 206,005 |  |  | 16,434 |  |  | 53,586 |  |  | 74,421 |  |  | 61,564 |  |  |
| Number of individuals | 11,548 |  |  | 1,164 |  |  | 3,387 |  |  | 4,060 |  |  | 2,937 |  |  |

Note. Adversity experiences and worries variables are weighted to the proportions of gender, age, ethnicity, country, and education obtained from the Office for National Statistics. Individual adversity experiences and worries variables are binary. Analyses were further adjusted for day of the week and time since lockdown began. OR = odds ratio. CI = confidence interval.

**Table S7.** Fixed-effects logistic regression models predicting within-individual change in self-harm behaviours from individual categories of adversity experiences and worries accounting for anxiety symptoms

| Self-harm behaviours |  |  |  |  |  |  |  |  |  |  |  |  |  |  |  |
| --- | --- | --- | --- | --- | --- | --- | --- | --- | --- | --- | --- | --- | --- | --- | --- |
|  | Total sample |  |  | Ages 18-29 |  |  | Ages 30-44 |  |  | Ages 45-59 |  |  | Ages 60+ |  |  |
| Variable | OR | 95% CI lower | 95% CI upper | OR | 95% CI lower | 95% CI upper | OR | 95% CI lower | 95% CI upper | OR | 95% CI lower | 95% CI upper | OR | 95% CI lower | 95% CI upper |
| Anxiety symptoms | 1.18 | 1.17 | 1.19 | 1.16 | 1.14 | 1.18 | 1.19 | 1.17 | 1.21 | 1.19 | 1.17 | 1.21 | 1.21 | 1.19 | 1.23 |
| Adversity experiences |  |  |  |  |  |  |  |  |  |  |  |  |  |  |  |
| Financial | 0.95 | 0.87 | 1.04 | 1.03 | 0.86 | 1.24 | 0.76 | 0.62 | 0.92 | 1.03 | 0.88 | 1.20 | 0.90 | 0.73 | 1.12 |
| Accessing essentials | 1.15 | 1.03 | 1.28 | 1.04 | 0.84 | 1.30 | 0.93 | 0.73 | 1.18 | 1.26 | 1.05 | 1.52 | 1.47 | 1.17 | 1.84 |
| COVID-19 illness | 1.35 | 1.20 | 1.53 | 2.77 | 2.07 | 3.72 | 1.00 | 0.78 | 1.29 | 1.46 | 1.20 | 1.78 | 0.92 | 0.69 | 1.22 |
| Illness of others/bereavement | 1.11 | 1.01 | 1.24 | 1.39 | 1.12 | 1.71 | 1.10 | 0.86 | 1.40 | 1.06 | 0.89 | 1.27 | 1.00 | 0.81 | 1.24 |
| Physical/psychological abuse | 4.14 | 3.82 | 4.49 | 3.09 | 2.58 | 3.71 | 6.33 | 5.28 | 7.60 | 3.54 | 3.08 | 4.07 | 4.54 | 3.85 | 5.35 |
| Worries |  |  |  |  |  |  |  |  |  |  |  |  |  |  |  |
| Financial | 1.08 | 1.00 | 1.17 | 0.96 | 0.82 | 1.12 | 1.09 | 0.92 | 1.29 | 1.24 | 1.07 | 1.43 | 1.05 | 0.88 | 1.25 |
| Accessing essentials | 1.17 | 1.08 | 1.26 | 1.41 | 1.22 | 1.64 | 1.02 | 0.87 | 1.19 | 1.11 | 0.97 | 1.28 | 1.12 | 0.96 | 1.31 |
| COVID-19 illness | 0.91 | 0.84 | 0.98 | 1.00 | 0.85 | 1.18 | 0.73 | 0.62 | 0.86 | 0.93 | 0.81 | 1.07 | 1.00 | 0.85 | 1.19 |
| Social/relationship concerns | 1.13 | 1.04 | 1.22 | 1.52 | 1.29 | 1.79 | 0.97 | 0.82 | 1.14 | 1.16 | 1.00 | 1.34 | 0.96 | 0.80 | 1.15 |
| Threats to safety | 1.07 | 0.99 | 1.15 | 0.98 | 0.83 | 1.15 | 1.47 | 1.24 | 1.74 | 1.03 | 0.90 | 1.18 | 0.96 | 0.82 | 1.13 |
| Number of observations | 63,383 |  |  | 7,654 |  |  | 18,540 |  |  | 23,707 |  |  | 13,482 |  |  |
| Number of individuals | 3,728 |  |  | 527 |  |  | 1,197 |  |  | 1,322 |  |  | 682 |  |  |

Note. Adversity experiences and worries variables are weighted to the proportions of gender, age, ethnicity, country, and education obtained from the Office for National Statistics. Individual adversity experiences and worries variables are binary. Analyses were further adjusted for day of the week and time since lockdown began. OR = odds ratio. CI = confidence interval.

**Table S8.** Fixed-effects logistic regression models predicting within-individual change in self-harm thoughts from individual categories of adversity experiences and worries accounting for depressive symptoms

| Self-harm thoughts |  |  |  |  |  |  |  |  |  |  |  |  |  |  |  |
| --- | --- | --- | --- | --- | --- | --- | --- | --- | --- | --- | --- | --- | --- | --- | --- |
|  | Total sample |  |  | Ages 18-29 |  |  | Ages 30-44 |  |  | Ages 45-59 |  |  | Ages 60+ |  |  |
| Variable | OR | 95%<br>CI<br>lower | 95%<br>CI<br>upper | OR | 95%<br>CI<br>lower | 95%<br>CI<br>upper | OR | 95%<br>CI<br>lower | 95%<br>CI<br>upper | OR | 95%<br>CI<br>lower | 95%<br>CI<br>upper | OR | 95%<br>CI<br>lower | 95%<br>CI<br>upper |
| Depressive symptoms | 1.37 | 1.37 | 1.38 | 1.35 | 1.33 | 1.36 | 1.37 | 1.36 | 1.38 | 1.38 | 1.37 | 1.39 | 1.39 | 1.38 | 1.41 |
| Adversity experiences |  |  |  |  |  |  |  |  |  |  |  |  |  |  |  |
| Financial | 1.00 | 0.94 | 1.05 | 1.06 | 0.93 | 1.21 | 0.97 | 0.87 | 1.09 | 0.96 | 0.87 | 1.05 | 1.06 | 0.94 | 1.20 |
| Accessing essentials | 1.13 | 1.04 | 1.22 | 0.77 | 0.61 | 0.96 | 0.84 | 0.71 | 0.99 | 1.51 | 1.31 | 1.74 | 1.22 | 1.05 | 1.41 |
| COVID-19 illness | 1.05 | 0.97 | 1.13 | 1.29 | 1.06 | 1.57 | 0.91 | 0.78 | 1.06 | 1.14 | 1.00 | 1.30 | 0.97 | 0.83 | 1.12 |
| Illness of others/<br>bereavement | 0.97 | 0.90 | 1.03 | 1.01 | 0.85 | 1.20 | 0.88 | 0.76 | 1.02 | 1.05 | 0.93 | 1.17 | 0.94 | 0.84 | 1.06 |
| Physical/ psychological<br>abuse | 2.65 | 2.50 | 2.81 | 3.07 | 2.60 | 3.63 | 2.77 | 2.43 | 3.16 | 2.59 | 2.34 | 2.88 | 2.50 | 2.26 | 2.78 |
| Worries |  |  |  |  |  |  |  |  |  |  |  |  |  |  |  |
| Financial | 1.18 | 1.13 | 1.24 | 1.15 | 1.03 | 1.27 | 1.05 | 0.96 | 1.15 | 1.30 | 1.19 | 1.42 | 1.20 | 1.09 | 1.31 |
| Accessing essentials | 1.01 | 0.96 | 1.05 | 1.23 | 1.09 | 1.38 | 0.90 | 0.82 | 0.99 | 0.84 | 0.77 | 0.92 | 1.16 | 1.06 | 1.26 |
| COVID-19 illness | 0.90 | 0.86 | 0.94 | 0.83 | 0.75 | 0.93 | 0.80 | 0.73 | 0.87 | 0.90 | 0.83 | 0.98 | 1.01 | 0.92 | 1.10 |
| Social/ relationship concerns | 1.32 | 1.26 | 1.38 | 1.58 | 1.41 | 1.77 | 1.31 | 1.19 | 1.43 | 1.32 | 1.21 | 1.44 | 1.23 | 1.13 | 1.34 |
| Threats to safety | 1.16 | 1.11 | 1.22 | 1.47 | 1.30 | 1.67 | 1.10 | 1.00 | 1.22 | 1.16 | 1.06 | 1.27 | 1.09 | 1.00 | 1.19 |
| Number of observations | 206,714 |  |  | 16,495 |  |  | 53,752 |  |  | 74,635 |  |  | 61,832 |  |  |
| Number of individuals | 11,580 |  |  | 1,168 |  |  | 3,394 |  |  | 4,069 |  |  | 2,949 |  |  |

Note. Adversity experiences and worries variables are weighted to the proportions of gender, age, ethnicity, country, and education obtained from the Office for National Statistics. Individual adversity experiences and worries variables are binary. Analyses were further adjusted for day of the week and time since lockdown began. OR = odds ratio. CI = confidence interval.

**Table S9.** Fixed-effects logistic regression models predicting within-individual change in self-harm behaviours from individual categories of adversity experiences and worries accounting for depressive symptoms

| Self-harm behaviours |  |  |  |  |  |  |  |  |  |  |  |  |  |  |  |
| --- | --- | --- | --- | --- | --- | --- | --- | --- | --- | --- | --- | --- | --- | --- | --- |
|  | Total sample |  |  | Ages 18-29 |  |  | Ages 30-44 |  |  | Ages 45-59 |  |  | Ages 60+ |  |  |
| Variable | OR | 95%<br>CI<br>lower | 95%<br>CI<br>upper | OR | 95%<br>CI<br>lower | 95%<br>CI<br>upper | OR | 95%<br>CI<br>lower | 95%<br>CI<br>upper | OR | 95%<br>CI<br>lower | 95%<br>CI<br>upper | OR | 95%<br>CI<br>lower | 95%<br>CI<br>upper |
| Depressive symptoms | 1.20 | 1.19 | 1.21 | 1.18 | 1.16 | 1.20 | 1.21 | 1.19 | 1.23 | 1.20 | 1.19 | 1.22 | 1.21 | 1.19 | 1.23 |
| Adversity experiences |  |  |  |  |  |  |  |  |  |  |  |  |  |  |  |
| Financial | 0.92 | 0.83 | 1.00 | 0.96 | 0.80 | 1.16 | 0.76 | 0.62 | 0.93 | 0.97 | 0.83 | 1.12 | 0.95 | 0.76 | 1.18 |
| Accessing essentials | 1.09 | 0.97 | 1.21 | 0.97 | 0.78 | 1.21 | 0.85 | 0.67 | 1.09 | 1.22 | 1.01 | 1.48 | 1.39 | 1.10 | 1.75 |
| COVID-19 illness | 1.32 | 1.17 | 1.50 | 2.99 | 2.20 | 4.05 | 0.88 | 0.68 | 1.14 | 1.49 | 1.22 | 1.82 | 0.91 | 0.68 | 1.20 |
| Illness of others/bereavement | 1.12 | 1.01 | 1.24 | 1.44 | 1.17 | 1.77 | 1.09 | 0.85 | 1.39 | 1.03 | 0.87 | 1.23 | 1.06 | 0.85 | 1.31 |
| Physical/ psychological abuse | 3.92 | 3.62 | 4.26 | 2.96 | 2.47 | 3.55 | 5.84 | 4.86 | 7.02 | 3.41 | 2.96 | 3.90 | 4.30 | 3.65 | 5.07 |
| Worries |  |  |  |  |  |  |  |  |  |  |  |  |  |  |  |
| Financial | 1.09 | 1.00 | 1.18 | 1.01 | 0.87 | 1.17 | 1.11 | 0.94 | 1.31 | 1.23 | 1.07 | 1.42 | 1.01 | 0.85 | 1.21 |
| Accessing essentials | 1.14 | 1.05 | 1.22 | 1.40 | 1.20 | 1.63 | 0.99 | 0.84 | 1.16 | 1.10 | 0.96 | 1.26 | 1.05 | 0.90 | 1.23 |
| COVID-19 illness | 0.95 | 0.88 | 1.03 | 1.07 | 0.91 | 1.26 | 0.76 | 0.65 | 0.90 | 0.95 | 0.83 | 1.09 | 1.06 | 0.90 | 1.26 |
| Social/ relationship concerns | 1.14 | 1.05 | 1.24 | 1.57 | 1.33 | 1.86 | 0.95 | 0.80 | 1.12 | 1.18 | 1.02 | 1.36 | 0.94 | 0.78 | 1.13 |
| Threats to safety | 1.09 | 1.01 | 1.18 | 1.01 | 0.85 | 1.19 | 1.49 | 1.26 | 1.77 | 1.08 | 0.94 | 1.23 | 0.95 | 0.81 | 1.12 |
| Number of observations | 63,767 |  |  | 7,687 |  |  | 18,594 |  |  | 23,859 |  |  | 13,627 |  |  |
| Number of individuals | 3,747 |  |  | 530 |  |  | 1,201 |  |  | 1,329 |  |  | 687 |  |  |

Note. Adversity experiences and worries variables are weighted to the proportions of gender, age, ethnicity, country, and education obtained from the Office for National Statistics. Individual adversity experiences and worries variables are binary. Analyses were further adjusted for day of the week and time since lockdown began. OR = odds ratio. CI = confidence interval.

**Table S10.** Fixed-effects logistic regression models predicting within-individual change in self-harm thoughts from individual categories of adversity experiences and worries with physical abuse and psychological abuse as individual adversity experiences

| Self-harm thoughts |  |  |  |  |  |  |  |  |  |  |  |  |  |  |  |
| --- | --- | --- | --- | --- | --- | --- | --- | --- | --- | --- | --- | --- | --- | --- | --- |
|  | Total sample |  |  | Ages 18-29 |  |  | Ages 30-44 |  |  | Ages 45-59 |  |  | Ages 60+ |  |  |
| Variable | OR | 95%<br>CI<br>lower | 95%<br>CI<br>upper | OR | 95%<br>CI<br>lower | 95%<br>CI<br>upper | OR | 95%<br>CI<br>lower | 95%<br>CI<br>upper | OR | 95%<br>CI<br>lower | 95%<br>CI<br>upper | OR | 95%<br>CI<br>lower | 95%<br>CI<br>upper |
| Adversity experiences |  |  |  |  |  |  |  |  |  |  |  |  |  |  |  |
| Financial | 1.18 | 1.13 | 1.24 | 1.26 | 1.12 | 1.42 | 1.16 | 1.04 | 1.28 | 1.12 | 1.03 | 1.22 | 1.25 | 1.12 | 1.40 |
| Accessing essentials | 1.28 | 1.19 | 1.38 | 0.94 | 0.77 | 1.15 | 1.01 | 0.87 | 1.17 | 1.66 | 1.46 | 1.89 | 1.34 | 1.17 | 1.53 |
| COVID-19 illness | 1.16 | 1.09 | 1.25 | 1.25 | 1.05 | 1.49 | 1.04 | 0.90 | 1.19 | 1.25 | 1.12 | 1.41 | 1.14 | 0.99 | 1.30 |
| Illness of others/bereavement | 1.15 | 1.08 | 1.22 | 1.29 | 1.11 | 1.51 | 1.02 | 0.89 | 1.16 | 1.23 | 1.11 | 1.37 | 1.09 | 0.98 | 1.21 |
| Physical abuse | 2.86 | 2.58 | 3.17 | 4.72 | 3.18 | 7.01 | 4.97 | 3.87 | 6.38 | 2.60 | 2.15 | 3.13 | 2.30 | 1.97 | 2.68 |
| Psychological abuse | 3.05 | 2.88 | 3.23 | 3.25 | 2.77 | 3.81 | 3.05 | 2.69 | 3.47 | 3.07 | 2.78 | 3.39 | 2.93 | 2.65 | 3.24 |
| Worries |  |  |  |  |  |  |  |  |  |  |  |  |  |  |  |
| Financial | 1.46 | 1.40 | 1.52 | 1.33 | 1.21 | 1.47 | 1.36 | 1.25 | 1.48 | 1.73 | 1.60 | 1.87 | 1.38 | 1.27 | 1.50 |
| Accessing essentials | 1.18 | 1.13 | 1.23 | 1.42 | 1.27 | 1.58 | 1.06 | 0.97 | 1.16 | 0.99 | 0.92 | 1.07 | 1.38 | 1.28 | 1.49 |
| COVID-19 illness | 0.94 | 0.91 | 0.98 | 0.82 | 0.75 | 0.91 | 0.80 | 0.74 | 0.87 | 0.97 | 0.90 | 1.05 | 1.13 | 1.05 | 1.22 |
| Social/relationship concerns | 1.63 | 1.56 | 1.69 | 1.92 | 1.73 | 2.14 | 1.69 | 1.55 | 1.83 | 1.62 | 1.51 | 1.75 | 1.44 | 1.33 | 1.56 |
| Threats to safety | 1.40 | 1.34 | 1.46 | 1.70 | 1.52 | 1.90 | 1.41 | 1.29 | 1.55 | 1.40 | 1.29 | 1.52 | 1.28 | 1.18 | 1.39 |
| Number of observations | 206,714 |  |  | 16,495 |  |  | 53,752 |  |  | 74,635 |  |  | 61,832 |  |  |
| Number of individuals | 11,580 |  |  | 1,168 |  |  | 3,394 |  |  | 4,069 |  |  | 2,949 |  |  |

Note. Adversity experiences and worries variables are weighted to the proportions of gender, age, ethnicity, country, and education obtained from the Office for National Statistics. Individual adversity experiences and worries variables are binary. Analyses were further adjusted for day of the week and time since lockdown began. OR = odds ratio. CI = confidence interval.

**Table S11.** Fixed-effects logistic regression models predicting within-individual change in self-harm behaviours from individual categories of adversity experiences and worries with physical abuse and psychological abuse as individual adversity experiences

| Self-harm behaviours |  |  |  |  |  |  |  |  |  |  |  |  |  |  |  |
| --- | --- | --- | --- | --- | --- | --- | --- | --- | --- | --- | --- | --- | --- | --- | --- |
|  | Total sample |  |  | Ages 18-29 |  |  | Ages 30-44 |  |  | Ages 45-59 |  |  | Ages 60+ |  |  |
| Variable | OR | 95%<br>CI<br>lower | 95%<br>CI<br>upper | OR | 95%<br>CI<br>lower | 95%<br>CI<br>upper | OR | 95%<br>CI<br>lower | 95%<br>CI<br>upper | OR | 95%<br>CI<br>lower | 95%<br>CI<br>upper | OR | 95%<br>CI<br>lower | 95%<br>CI<br>upper |
| Adversity experiences |  |  |  |  |  |  |  |  |  |  |  |  |  |  |  |
| Financial | 1.01 | 0.92 | 1.11 | 0.96 | 0.80 | 1.16 | 0.78 | 0.64 | 0.95 | 1.13 | 0.97 | 1.31 | 1.11 | 0.89 | 1.37 |
| Accessing essentials | 1.19 | 1.07 | 1.33 | 1.09 | 0.87 | 1.37 | 0.91 | 0.72 | 1.16 | 1.36 | 1.13 | 1.64 | 1.49 | 1.18 | 1.89 |
| COVID-19 illness | 1.33 | 1.17 | 1.50 | 2.33 | 1.74 | 3.12 | 1.00 | 0.78 | 1.30 | 1.44 | 1.19 | 1.75 | 1.01 | 0.76 | 1.33 |
| Illness of others/bereavement | 1.22 | 1.10 | 1.35 | 1.54 | 1.24 | 1.91 | 1.21 | 0.95 | 1.54 | 1.11 | 0.93 | 1.33 | 1.14 | 0.92 | 1.40 |
| Physical abuse | 8.85 | 7.85 | 9.96 | 19.53 | 13.53 | 28.19 | 15.70 | 11.62 | 21.21 | 5.77 | 4.74 | 7.02 | 7.83 | 6.36 | 9.63 |
| Psychological abuse | 2.39 | 2.19 | 2.61 | 1.74 | 1.44 | 2.10 | 2.54 | 2.08 | 3.11 | 2.69 | 2.32 | 3.12 | 2.55 | 2.12 | 3.06 |
| Worries |  |  |  |  |  |  |  |  |  |  |  |  |  |  |  |
| Financial | 1.24 | 1.15 | 1.34 | 1.16 | 1.00 | 1.35 | 1.34 | 1.14 | 1.58 | 1.42 | 1.24 | 1.64 | 1.08 | 0.91 | 1.28 |
| Accessing essentials | 1.24 | 1.15 | 1.34 | 1.61 | 1.38 | 1.87 | 1.11 | 0.95 | 1.30 | 1.17 | 1.02 | 1.34 | 1.14 | 0.98 | 1.34 |
| COVID-19 illness | 0.97 | 0.90 | 1.04 | 1.06 | 0.90 | 1.24 | 0.77 | 0.65 | 0.91 | 0.95 | 0.83 | 1.08 | 1.16 | 0.98 | 1.36 |
| Social/relationship concerns | 1.30 | 1.20 | 1.41 | 1.57 | 1.32 | 1.85 | 1.18 | 1.00 | 1.40 | 1.33 | 1.16 | 1.53 | 1.13 | 0.94 | 1.35 |
| Threats to safety | 1.22 | 1.13 | 1.32 | 1.11 | 0.94 | 1.31 | 1.72 | 1.46 | 2.04 | 1.21 | 1.06 | 1.38 | 1.04 | 0.88 | 1.22 |
| Number of observations | 63,767 |  |  | 7,687 |  |  | 18,594 |  |  | 23,859 |  |  | 13,627 |  |  |
| Number of individuals | 3,747 |  |  | 530 |  |  | 1,201 |  |  | 1,329 |  |  | 687 |  |  |

Note. Adversity experiences and worries variables are weighted to the proportions of gender, age, ethnicity, country, and education obtained from the Office for National Statistics. Individual adversity experiences and worries variables are binary. Analyses were further adjusted for day of the week and time since lockdown began. OR = odds ratio. CI = confidence interval.

